# Facilitators and Barriers for Telemedicine Systems in India from Multiple Stakeholder Perspectives and Settings

**DOI:** 10.1101/2023.04.23.23288980

**Authors:** Aparna Venkataraman, Najiya Fatma, Sisira Edirippulige, Varun Ramamohan

## Abstract

Telemedicine is viewed as a crucial tool for addressing the challenges of limited medical resources at healthcare facilities. However, its adoption in healthcare is not entirely realised due to perceived barriers. This systematic review outlines the critical facilitators and barriers that influence the implementation of telemedicine in the Indian healthcare system, observed at the infrastructural, socio-cultural, regulatory and financial levels, from the perspectives of healthcare providers, patients, patient caregivers, society, health organisations and the government. This review complies with the current PRISMA-P protocol. 1200 peer-reviewed studies published from December 2016 to December 2021 in the PubMed, Cochrane, Scopus, Web of Science, CINAHL, MEDLINE and PsycInfo databases were considered for the title and abstract screening, after which 157 articles were chosen for the full-text review. In the end, 26 studies were selected for data synthesis. Data privacy and security concerns, doctor and patient resistance to information and communications technology (ICT), poor infrastructure, and lack of ICT training were considered significant barriers to implementing telemedicine. However, reduced healthcare delivery costs, improved patient access to healthcare in remote areas, and reduced patient waiting times all helped promote telemedicine implementation. The review outcomes also revealed that the barriers and facilitators at the regulatory and financial level largely influenced the adoption of telemedicine systems in India, with 59% (n=20) articles citing a reduction in healthcare delivery costs as the critical facilitator and 59% citing fear of violation of patient privacy and security as the significant barrier.

## 1. Introduction

Today, one of India’s most challenging problems is delivering affordable healthcare to its population ^1^. In 2019, around 66% of the Indian population lived in rural areas ^2,3^, and there was a severe lack of 17,459 specialists, 3,184 radiographers, 1,484 doctors, 6,412 nursing staff, 6,743 auxiliary nurse midwives, and 12,065 lab technicians in the rural public health system ^4^. However, reforming the Indian healthcare system is challenging due to considerable inequities in medical resource distribution across rural and urban parts of India. There are no simple solutions to India’s healthcare problems; nevertheless, harnessing India’s developments in information and communications technology (ICT) in healthcare delivery through telemedicine presents one potential approach towards reducing these healthcare delivery and quality disparities.

WHO has defined telemedicine as “the delivery of health care services, where distance is a critical factor, by all health care professionals using information and communication technologies for the exchange of valid information for diagnosis, treatment and prevention of disease and injuries, research and evaluation, and for the continuing education of health care providers, all in the interests of advancing the health of individuals and their communities” ^5^. Telemedicine-based consultation has been shown to diagnose and treat diseases at lower healthcare delivery costs ^6^.

In 2000, the Indian government adopted telemedicine for healthcare delivery. Kerala, Maharashtra, Punjab, and Tamil Nadu launched a telemedicine project in early 2000 to enhance rural healthcare access by connecting multispecialty hospitals to the Department of Information Technology (DIT). DIT connects several medical research centres and tertiary care hospitals, including AIIMS Delhi, SGPGIMS, and PGIMER Chandigarh ^7^. In 2001, The Indian Space Research Organisation (ISRO) connected Apollo Hospitals in Chennai and Chittoor to deliver telemedicine, and currently, 205 rural hospitals, 245 hospitals, and 40 super-specialty hospitals are part of ISRO’s telemedicine network. In recent years, the Indian government has initiated several new telemedicine projects, such as the National Cancer Network (ONCONET), the National Rural Telemedicine Network, the telemedicine initiative in North East India by the North Eastern Space Applications Centre, and the Integrated Disease Surveillance Project ^8^. Additionally, some private hospitals, such as Apollo Hospitals, Amrita Institute of Medical Sciences, Asia Heart Foundation, Aravind Eye Care, Escorts Heart Institute, and Narayana Hrudayalaya, have also reported using telemedicine for healthcare ^9^. Further, in response to the COVID-19 pandemic, the Indian Ministry of Health and Family Welfare issued guidelines on March 25, 2020, allowing a registered medical practitioner (RMP) to practise telemedicine and defined the functions of RMPs, healthcare providers, and technological platforms to ensure the continued delivery of healthcare services to the public ^10,11^.

With regard to the literature associated with the practice of telemedicine in India, after the onset of COVID-19, the literature primarily comprised narrative articles about the telemedicine framework and the challenges in its implementation (Agarwal and Biswas ^12^; Dash, Aarthy ^13^; Garg, Gangadharan ^14^; Dinakaran, Manjunatha ^15^). A few studies also focused on the scope of the telemedicine framework for patient care in specific clinical specialties, such as neurology (Appireddy, Bendahan ^16^), abortion (Chandrasekaran, Chandrashekar ^17^), dentistry (Deshpande, Patil ^18^), child and adolescent healthcare (Galagali, Ghosh ^19^), sleep medicine (Gupta, Kumar ^20^), ophthalmology (Jayadev, Mahendradas ^21^; Sharma, Jain ^22^), dermatology (Pasquali, Sonthalia ^23^), psychiatry (Dinakaran, Basavarajappa ^25^; Vadlamani, Sharma ^26^), psychiatric rehabilitation (Jayarajan, Sivakumar ^27^), diabetes (Pradeepa, Rajalakshmi ^28^), and ICU Services (Ramakrishnan, Tirupakuzhi Vijayaraghavan ^29^) among others.

Additionally, some reviews have also reported the benefits and barriers to telemedicine implementation in India. Sharma and Prashar ^30^ summarised the merits and challenges of implementing eHealth systems in India pertaining to healthcare administration, finance, and healthcare delivery from multiple perspectives through a narrative literature review. Verma, Krishnan ^31^ systematically reviewed the literature on telemedicine systems in India and elaborated on the pitfalls and barriers that limit its utility among the general public and healthcare providers. Chandwani and Dwivedi ^32^ presented their viewpoints on the scope of telemedicine systems and the observed barriers at the policy, socio-cultural, and resources level that impacts its diffusion. However, none of these reviews discussed the barriers and facilitators to various modes of telemedicine that are observed at different healthcare delivery tiers and from multiple perspectives in the Indian healthcare system.

Most of the studies discussed here, and the others in the telemedicine literature, only reported the barriers and facilitators to a particular mode of telemedicine with regard to a specific clinic specialty in a single centre and corresponded to a homogeneous target population. Besides, the studies also did not analyse the perceptions of all stakeholders regarding the barriers and facilitators, which might have omitted certain factors that could have a significant impact on the adoption and acceptance of telemedicine, as the perceptions and preferences of all the stakeholders must be incorporated to implement effective telemedicine systems. Therefore, to the best of our knowledge, no published systematic review provides a comprehensive summary of the barriers and facilitators to various modes of telemedicine in the Indian healthcare system from multiple stakeholder perspectives. Thus, this review bridges this vital gap and contributes to the literature through a systematic review of the infrastructural, socio-cultural, regulatory and financial facilitators and barriers to various modes of telemedicine systems from multiple stakeholder perspectives and settings in the Indian context.

## 2. Methods

### 2.1 Protocol Registration

This review protocol is registered with PROSPERO International prospective register of systematic reviews (CRD42022306271) ^33^.

### 2.2 Review Framework

A rapid systematic review was conducted with the PICO (Population, Intervention, Comparison and Outcomes) framework ^34^ to assess the recent facilitators and barriers influencing the implementation of telemedicine systems in India.

#### Nature of Population

Studies in which the participants undertook monitoring/screening/diagnostic tests/treatment in telemedicine-based intervention in India’s primary, secondary, and tertiary healthcare systems were included. The viewpoints of telemedicine providers or experts in Indian medical centres or hospitals were also included in the review.

#### Type of Intervention

All studies that considered the provision of healthcare via telemedicine, regardless of any specific intervention or clinical specialty, were included.

#### Comparator System

The obstructors and facilitators for the adoption of the telemedicine framework within the traditional healthcare system in India were analysed.

#### Review Outcomes

The review’s primary outcomes are the recent facilitators and barriers to adopting telemedicine systems in India from the perspectives of healthcare providers, patients, patient caregivers, society, health organisations and government. In this review’s context, the factors that helped the relevant stakeholders (e.g., patients or providers) to adopt telemedicine systems are referred to as facilitators, and the factors that obstructed stakeholders from adopting telemedicine are referred to as barriers.

The authors aim to address the following research questions (RQ) through this systematic review:

RQ1: What are the critical infrastructure, socio-cultural, regulatory and financial facilitators, and barriers to implementing telemedicine in Indian healthcare facilities?

RQ2: What are the significant facilitators and barriers to different modes of telemedicine consultation in primary, secondary, and tertiary healthcare sectors from multiple stakeholder perspectives?

### 2.3 Search Strategy

Seven digital databases – PubMed, Cochrane, Scopus, Web of Science, CINAHL, MEDLINE and PsycInfo were selected for retrieving studies using six groups of keywords and all their relevant words comprising MeSH (Medical Subject Headings) and title and text keywords. Different combinations of keyword groups were assessed, and the best combination that retrieved the maximum number of relevant studies from the databases was selected for further review. Additionally, the following restrictions were also included to refine search outcomes from the databases,

*Articles in English; Articles published during the review period (2016 December to 2021 December); Peer-reviewed articles; Articles with full-text availability; Articles dealing with human population; Search with different variations of the keywords*

All the restrictions were not added at once; only those relevant to a specific database were included in the search query. The literature search of chosen databases was conducted between December 2021 and February 2022, and 1726 articles published during the review period were selected. Furthermore, references from all selected studies were also screened to gather additional publications that the search query may have missed.

The keyword groups, database search queries, and search results from the databases for the review period have been included in the supplementary file1.

### 2.4 Study Screening

The initial search yielded 1726 articles in total. Following de-duplication, 526 duplicate articles were removed, and 1200 studies remained for further review. One author (AV) reviewed the titles and abstracts of the publications for inclusion. Following the title and abstract screening, 1043 articles were excluded owing to irrelevance to this review, leaving 157 for the next step. In the second screening step, two authors (AV and NF) examined these articles’ titles, abstracts, and contents to determine their relevance to the research question, narrowing the results to 59.

### 2.5 Study Eligibility Criteria

The inclusion and exclusion criteria were then applied to the remaining 59 articles. The selection procedure is illustrated as a PRISMA (Preferred Reporting Items for Systematic Reviews and Meta-Analyses) flowchart ^35^ in Figure 1.

**Figure 1:**
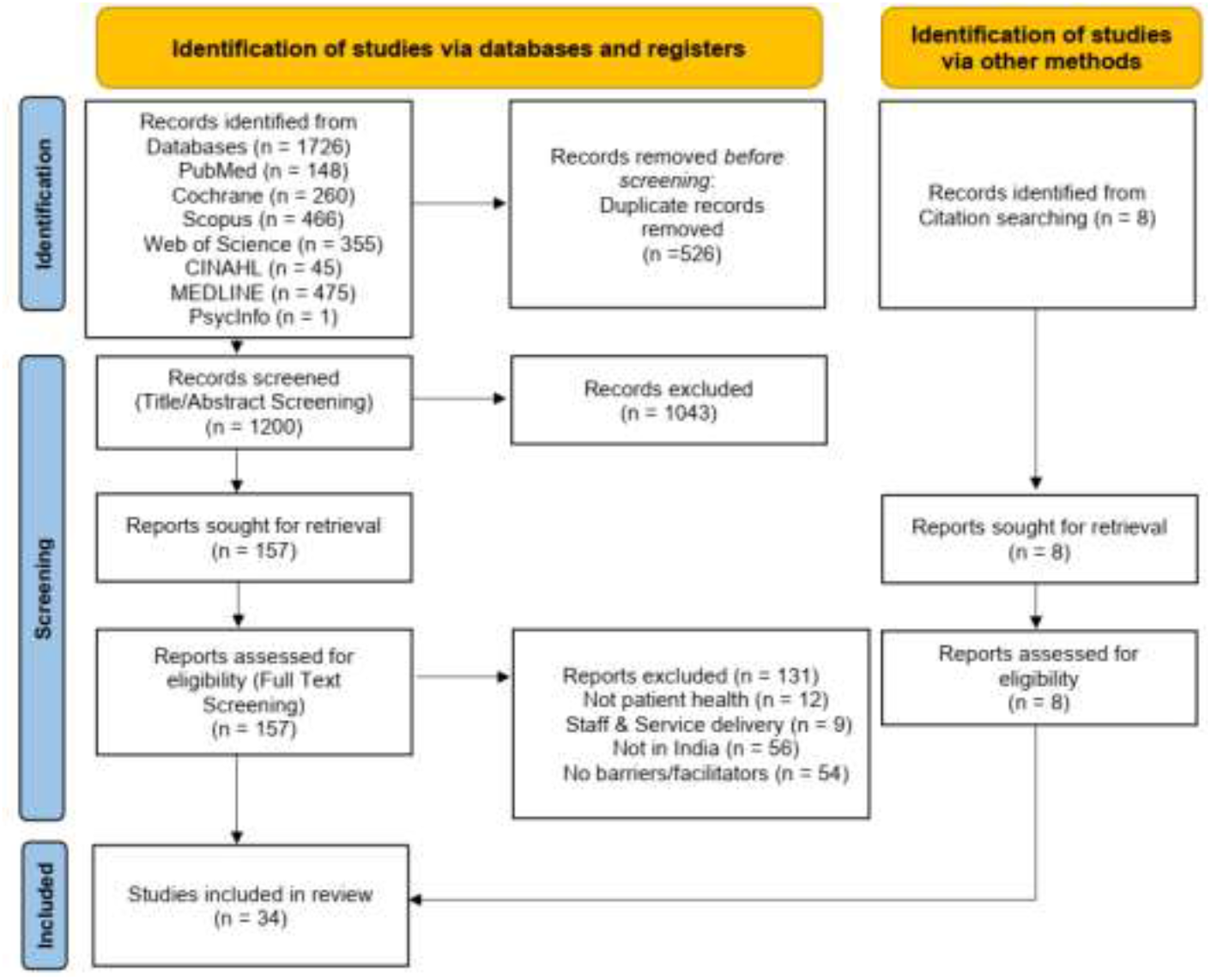
Article selection procedure flowchart.

#### Inclusion Criteria

Articles with the following attributes were included in the review:

1. Peer-reviewed articles in English with a full text published between December 2016 to December 2021.
2. Articles with telemedicine intervention dealing with patient health in any clinical specialty in India ^36,37^.
3. Reviews and narrative articles including opinions of healthcare providers/patients/health experts in a medical centre/hospital in India ^22,31^.

#### Exclusion Criteria

The following types of articles were excluded:

1. Articles not directly associated with patient health but with staff training, health education, medical administration, medical records, digital health, health information systems, and telemedicine framework ^38^.
2. Articles with telemedicine interventions not concerning patient healthcare services such as monitoring/screening/diagnostic tests/treatment but only dealing with communication among staff and service delivery. However, if the articles focussed on telemedicine interventions for patient healthcare along with some of the excluded areas in (a) and (b), then those articles were considered for full-text screening ^39^.
3. Articles about barriers or facilitators for telemedicine systems not based in India ^40^.
4. If research outcomes of articles did not address barriers or facilitators to adopting telemedicine systems ^41^.

Two authors (AV and NF) independently analysed the full text of 59 articles obtained after the second screening process. Any unresolved differences of opinion between the authors regarding the selection were resolved through discussions in meetings until a consensus was reached. In the end, 26 articles were finalised for data synthesis. Additionally, 8 more articles were also included for review from the search of references of all the selected studies.

The 131 rejected studies and their exclusion criteria are outlined in Supplementary File 2.

## 3. Results

The selected articles were summarised by outlining the study participants, study perspective, healthcare system tier, clinical specialty, mode of delivery of telemedicine intervention, and the facilitators and barriers to implementation, which have been included in the supplementary file1.

### 3.1 Study perspectives and survey population demographics

Out of the 34 selected studies, six outlined the barriers and facilitators from doctors’ views, and one outlined the perspectives of health workers and other technical staff. Likewise, six studies outlined the perspectives of patients, and two studies dealt with the viewpoints of patients and their caregivers. Furthermore, seven studies narrated the most observed barriers and facilitators, which were considered the viewpoints of the society in this review. The remaining twelve articles summarised the barriers and facilitators from multiple perspectives, which included healthcare providers, patients, patient caregivers, society, health organisations and government. The interventions in the selected studies were implemented in several North Indian, South Indian, and West Indian states. Most of the studies were conducted in North Indian states and Karnataka, and none of the telemedicine interventions are from the Eastern states of India.

A total of 18 studies interpreted the barriers and facilitators for telemedicine interventions through primary data gathered from the surveys of healthcare providers and patients. Among these, six studies collected data exclusively from physicians, healthcare providers, and other technical staff. Likewise, eight studies utilised data collected through surveys of patients and their caregivers only, and the remaining four articles collected data from a mixed group of respondents.

From Table I, it can be observed that most study participants were male and aged between 18 and 67 years. Most patients belonged to the lower or middle socioeconomic class (modified Kuppuswamy’s class ^42^), with approximately 60% from rural regions. Patient caregivers were primarily females belonging to the age group of 26 to 52 years. Likewise, the data collected from doctors, other healthcare providers, and technical staff in the articles showed that the doctors aged between 34 to 45 years with an experience of 8 to 12 years. Resident doctors below 30 years of age had an experience of fewer than five years. Similarly, other healthcare providers and technical staff were aged 25 to 56 years with an experience of 3 to 7 years. Doctors, healthcare providers, and technical staff practised various clinical specialties in different clinical settings.

**Table I:**
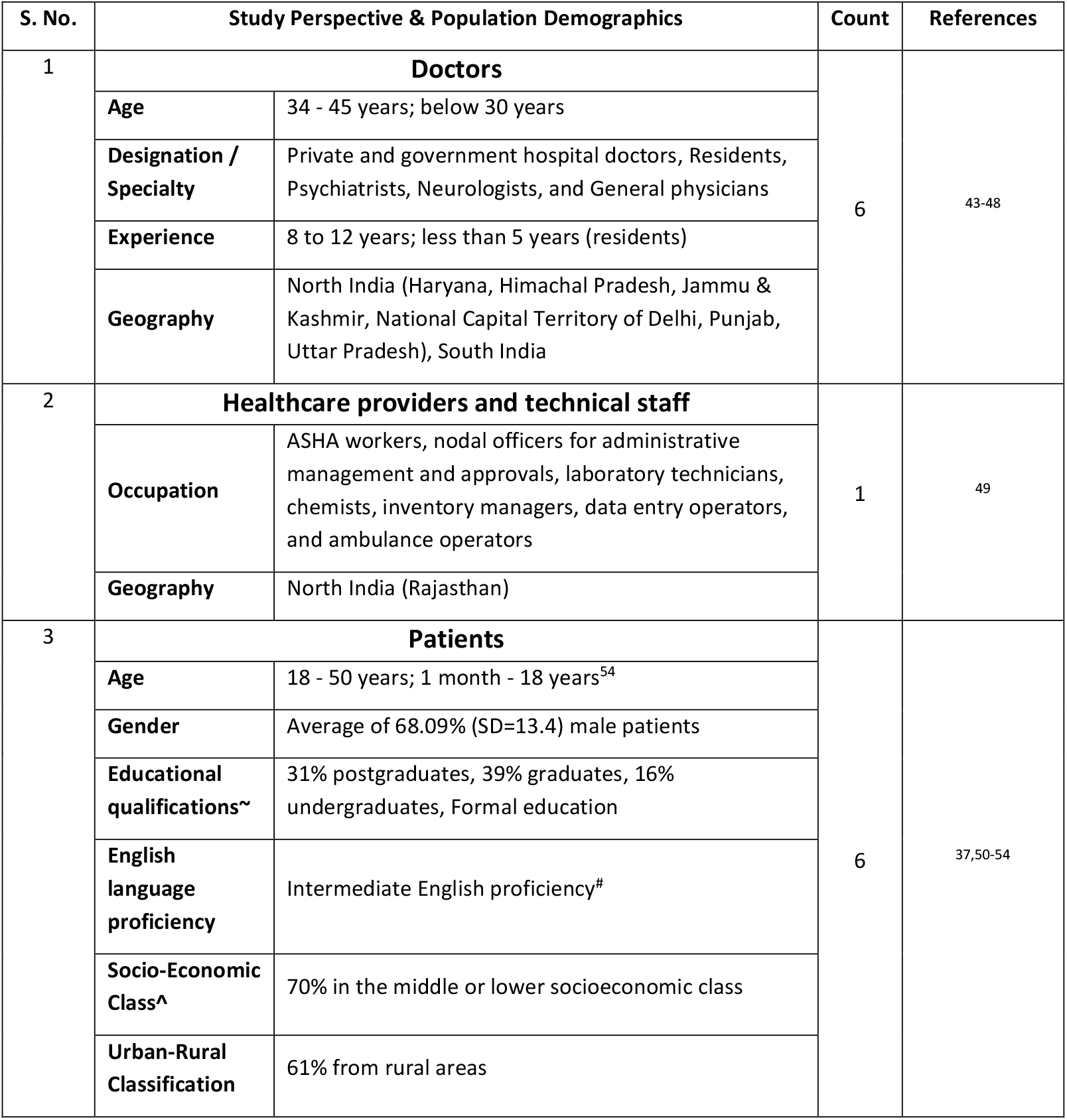
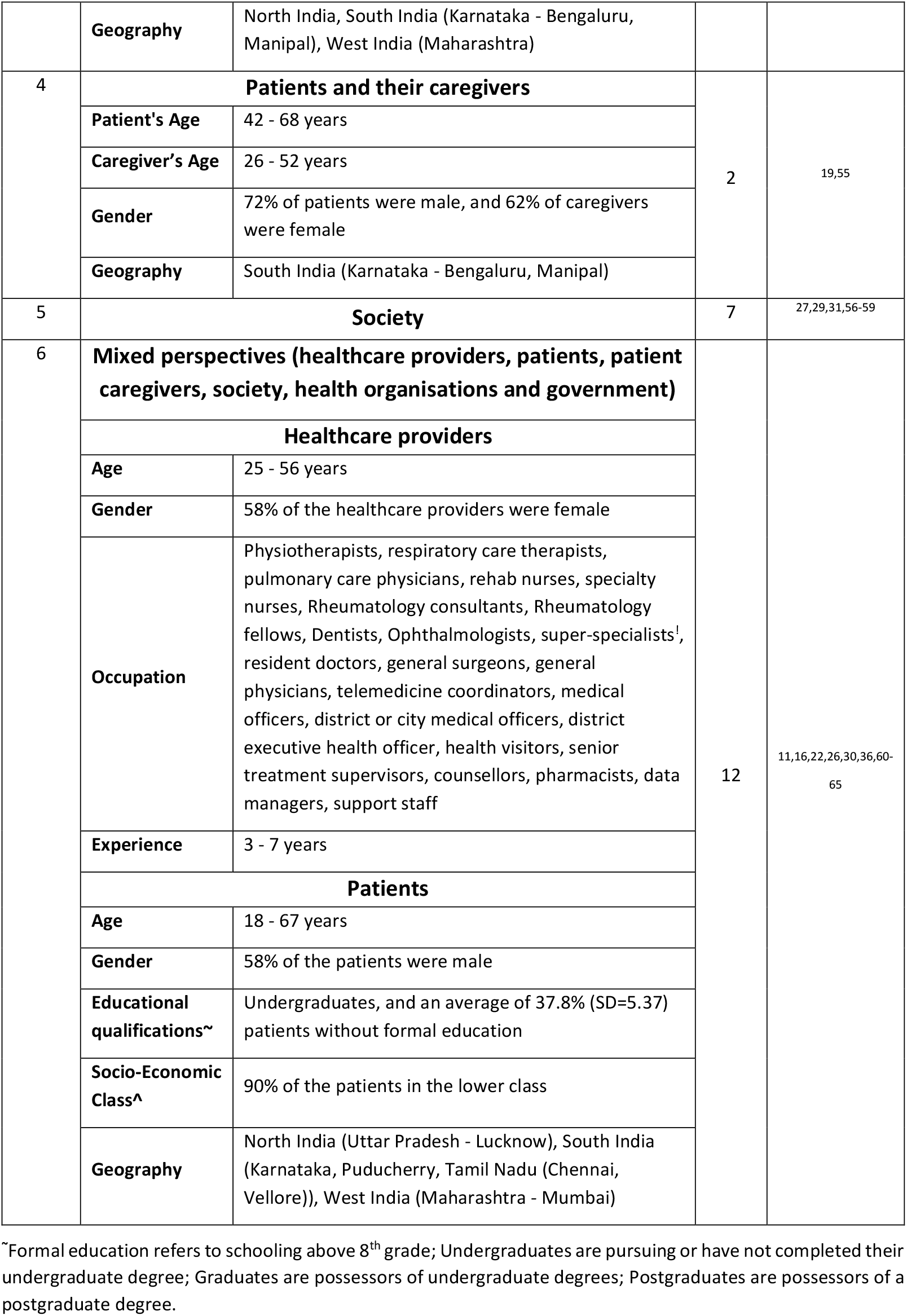
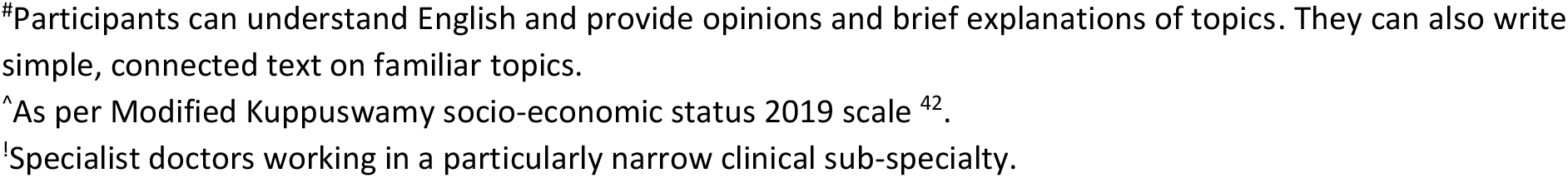
Study perspectives and population demographics in the selected articles.

### 3.2 Clinical specialty and healthcare sector

Out of 34 articles, seventeen focused on primary healthcare, ten on secondary and seven on tertiary care, as indicated in Table II. The clinical specialties observed in the articles included teleneurology (n=1), telemedicine for stroke management (n=1), telerehabilitation (including stroke rehabilitation) (n=3), teledentistry (n=1), teleurology (n=1), telepsychiatry (including geriatric psychiatry) (n=4), dementia care (n=1), telerheumatology (n=1), tuberculosis care (n=2), tele-ICU (n=1), teleophthalmology (n=2), telepaediatrics (including paediatric HIV care, child and adolescent healthcare) (n=3), telehepatology (n=1), and surgical endocrinology (n=1). One article focussed on triaging using telemedicine. The other ten articles did not emphasise any clinical speciality and outlined the barriers and facilitators to all telemedicine interventions.

**Table II:** Clinical Specialties and healthcare sectors addressed in the selected studies.

| Healthcare Sector | Clinical Speciality | Number of articles | References |
| --- | --- | --- | --- |
| Primary | Dementia care | 1 | 44 |
|  | Telepsychiatry | 4 | 26,51,56,61 |
|  | Telemedicine | 10 | 30,31,43,46-50,57,58 |
|  | Tuberculosis care | 1 | 52 |
|  | Triaging | 1 | 65 |
| Secondary | Teleneurology | 1 | 16 |
|  | Telerehabilitation | 1 | 36 |
|  | Telepaediatrics | 3 | 11,19,54 |
|  | Telemedicine for stroke management | 1 | 45 |
|  | Teledentistry | 1 | 60 |
|  | Teleophthalmology | 2 | 22,59 |
|  | Tuberculosis care | 1 | 63 |
| Tertiary | Teleurology | 1 | 37 |
|  | Telerehabilitation | 2 | 27,55 |
|  | Telerheumatology | 1 | 62 |
|  | Tele-ICU | 1 | 29 |
|  | Telehepatology | 1 | 53 |
|  | Surgical endocrinology | 1 | 64 |

### 3.3 Mode of delivery of telemedicine interventions

The Indian telemedicine framework permits the exchange of medical information for telemedicine consultations via multiple real-time and asynchronous communication media and channels, such as text communication (e.g., SMS, email), audio, and video. This includes messaging applications, websites set up expressly for telemedicine care, phone calls, video on chat platforms, email, and fax ^10^. In this review, the modes of telemedicine delivery in the articles were categorised into Tele/Video and mHealth, as shown in Table III. Mobile health (m-Health) refers to the use of mobile phones for healthcare delivery and is further categorised into mobile teleconsultation and non-smartphone-based technology in this review. Similarly, Tele/Video is further classified into video teleconferencing and teleconsultation modes.

**Table III:** Modes of telemedicine delivery in the review articles.

| Mode of Delivery | Type | Count | References |
| --- | --- | --- | --- |
| Tele/Video | Teleconsultation | 7 | 27,44,47,50,56,60,61 |
|  | Video teleconferencing | 5 | 16,19,26,54,64 |
| mHealth | Mobile teleconsultation | 9 | 22,36,37,45,52,53,55,62,65 |
|  | Non-smartphone-based technology | 1 | 63 |

The definition adopted for classifying the selected articles to these delivery modes is elucidated as follows:

- Teleconsultation: A broad term representing one-on-one audio/video communication between primary physicians/medical specialists and patients through an ICT medium, introduced to account for studies where the delivery medium is not clearly specified.
- Video teleconferencing: Video communication between multiple stakeholders such as super-specialists, primary physicians, resident doctors, patients, patient caregivers and technical coordinators (at the hospital site).
- Mobile teleconsultation: Use of mobile phones for communication between healthcare providers and patients via video calls, voice calls, messages, video and audio instructions for patients, real-time sharing of medical data like photographs and prescriptions, and smartphone applications for delivery of healthcare.
- Non-smartphone-based technology: Use of cell phones with no internet connectivity for communication between healthcare providers and patients to deliver healthcare services via voice calls and messages.

A total of 22 articles disclosed their mode of telemedicine intervention, among which five articles focussed on video teleconferencing. A large proportion of articles (n=10), categorised as ‘mHealth’, highlighted the use of mobile phones for teleconsultation, among which nine articles utilised smartphones (two studies operated their unique mobile applications ^22,37^), and one study used non-smartphone-based technology in their telemedicine interventions. Seven studies that employed audio/video communication between providers and patients but did not disclose the delivery medium were categorised into ‘teleconsultation’ in this review. Furthermore, a total of ten narrative articles did not provide details regarding the mode of telemedicine intervention, and the remaining two studies dealt with barriers and facilitators for e-health systems in their narration.

### 3.4 Facilitators and barriers for telemedicine interventions

Among the 34 selected articles, 59% reported a reduction in healthcare delivery costs as one of the significant facilitators of telemedicine systems in India. Likewise, nearly 50% of the articles preferred telemedicine for follow-up consultations and highlighted telemedicine’s role in enhancing access to medical services in remote areas. Around 40% of the selected articles emphasised the importance of teleconsultations in saving time and reducing the frequency of hospital visits. Furthermore, 35% of the studies reported that telemedicine reduced the commute to hospitals for consultations, and 30% of the articles reported ease of consultation using telemedicine systems as one of its facilitators.

Likewise, 59% of the articles identified the fear of violating privacy and the security of patients’ medical information as the most significant barrier to adopting telemedicine systems at healthcare facilities in India. Patients’ preconceived doubts regarding the quality and efficacy of virtual consultation and their suitability as an alternate mode of healthcare delivery were the other barriers identified by 56% of the articles. Similarly, 47% of the articles reported ambiguities concerning the regulations in the Indian telemedicine framework and legal liabilities and lower technology literacy in rural and remote areas as crucial factors for poor adoption of telemedicine systems. Unavailability of technological resources and inadequate internet connectivity were also identified as barriers to the implementation of telemedicine systems by about 40% and 32% of the articles, respectively. Similarly, the lack of dedicated and trained workforces for telemedicine interventions and the requirement of physical assessments over virtual consultations in certain medical specialties were highlighted as barriers to telemedicine systems in 29% of the articles.

Furthermore, although 59% of the articles identified reduction in treatment costs as one of the significant facilitators of telemedicine systems, 35% of the studies reported that telemedicine systems were a financial burden to healthcare providers and patients. Therefore, the barriers and facilitators were categorised and analysed further by study perspective, health system tier, and mode of telemedicine delivery to understand the impact of various factors on the adoption of telemedicine in the Indian healthcare system.

A list of barriers and facilitators categorised by the articles in which they are mentioned is included in the supplementary file1.

## 4. Discussion - Categorisation of barriers and facilitators

The contents of the included articles were analysed to identify the significant factors that influence the implementation of telemedicine systems, and the facilitators and barriers were grouped into three categories of factors: (a) infrastructural factors, (b) socio-cultural factors, and (c) regulatory and financial factors.

In this review’s context, the factors that persuade policymakers and healthcare organisations to accept telemedicine as an effective approach to patient treatment and consequently finance telemedicine are the regulatory and financial factors. The factors influencing healthcare staff and patients to embrace the concept and technological skills necessary to access telemedicine are categorised as socio-cultural and infrastructural factors.

### 4.1 Infrastructural factors

The core infrastructure for telemedicine encompasses both technological and human resources. The function of technology resources in a telemedicine system is to act as an effective communication medium for human stakeholders to facilitate the transfer of medical information ^64^. The availability of infrastructure, professional knowledge, and training all impact the implementation of telemedicine in India. All the infrastructural barriers and facilitators are reported from the perspectives of doctors, health workers, technical staff, patients, and their caregivers.

#### Barriers

The unavailability of reliable internet connectivity and hardware infrastructure was reported as a critical barrier to telemedicine adoption in primary healthcare systems and all modes of real-time delivery ^26,30,31,47,49,50,52,58,61,65^. The concerns with internet quality (e.g., slow speeds, poor connectivity, and high costs) have been highlighted in the literature as a continuing challenge for the operation of telemedicine systems. Most telemedicine applications require a reliable internet connection for real-time teleconsultations. Due to the unavailability of high-speed internet connectivity in rural and remote locations, the likelihood of utilising telemedicine-based consultations was lower ^29^. Also, many state-of-the-art telemedicine equipments become obsolete as telemedicine advances, and the stakeholders cannot afford to replace the outdated technologies ^58^. The failure of the telemedicine network in Madhya Pradesh illustrates how ISRO-sponsored telemedicine equipment became obsolete and non-functional over time. The telemedicine network in the state collapsed as the government could not repair or replace the equipment due to the high cost of infrastructure ^66^.

Additionally, we also identified the lack of training and awareness regarding the usage of telemedicine and the associated equipment among patients and providers as a significant barrier to the adoption of telemedicine systems in primary and secondary healthcare ^26,30,31,36,44,45,49,56,58,61,65^. This may probably be due to the limited dissemination of information regarding telemedicine through workshops, conferences, seminars, and other platforms. A study ^50^ reported that this lack of technical awareness of telemedicine negatively impacts the patient’s willingness to adopt telemedicine. Few studies also reported that the patients expressed concern regarding the dearth of availability and expertise in telemedicine interventions ^36,58^.

#### Facilitators

The inherent benefits of employing telemedicine may be realised if the process is convenient for the practitioners and patients. Accordingly, studies ^16,44,50,52,62^ reported the factors related to convenience in consultation as the critical facilitators at this level in all three healthcare tiers. One of the significant facilitators of telemedicine systems is their ability to save time for healthcare providers and patients. Online consultation is also reported to increase convenience by minimising patient waiting time at specialist consultations and outpatient units. Telemedicine also reduces the travel burden on patients and their families by reducing the number of in-person visits and the associated costs.

Studies with video teleconferencing and mobile consultation interventions stated that telemedicine saves time, travel, and effort ^22,27,31,53,55,58,64^ in secondary ^11,19,22,59,60,63^ and tertiary care ^29,37,53,64^ and reduces the hospital visits where an in-person consultation is not warranted.

### 4.2 Socio-cultural factors

The socio-cultural factors relate to the prevailing conventions that influence health-seeking behaviour in a population. This research highlights various social and cultural factors that may impact telemedicine adoption in India. All the barriers and facilitators at the socio-cultural level are reported from the perspectives of doctors, health workers, technical staff, patients, and their caregivers.

#### Barriers

Patients’ preconceived doubts regarding the quality and efficacy of the virtual consultations provided by telemedicine and their suitability as an alternate mode of healthcare delivery were identified as one of the most critical barriers at the socio-cultural level ^29-31,36,47,56,58,63^. This was a significant barrier to the adoption of telemedicine in secondary care ^16,22,36,45,54,59,60,63^. As reported in ^22,45,54^, it appeared to be challenging for patients to accept that telemedicine can adequately replace in-person examinations. Patients preferred in-person consultations for various reasons, such as their conviction that the physical presence of a healthcare practitioner would lead to a better interpretation of body ailments or because they lacked confidence in performing a self-exam and reporting it as per the doctors’ recommendations or simply because it was their preference.

Even some healthcare providers thought that patient consultation and therapy should involve a physical examination and in-depth observation rather than remote consultation via telemedicine ^11,46,57,58^. This attitude towards telemedicine represents a potential barrier to further advancements ^36,37,52,53,56,64^. Healthcare providers acknowledged the technical ease of virtual consultations but expressed doubts about the quality of virtual examinations since the patients may not be technologically competent. The practitioners were also concerned that the patients’ temporary eyesight and hearing impairment might lead to erroneous virtual tests and improper compliance with instructions ^44^.

Another significant barrier at this level was the lack of ICT literacy and knowledge of its application in healthcare, particularly in primary healthcare. According to some doctors, this poor technological literacy creates resistance to adopting change ^43,58^ and poses a critical barrier to the acceptance and development of telemedicine. Meanwhile, some studies identified resistance to telemedicine implementation by doctors and patients as a cultural barrier ^11,19,30,31,36,44,49,58^. The end-user resistance to telemedicine could at least partially be attributed to their lack of technology-related knowledge, expertise, and experience, which can also affect their perceptions and willingness to adopt telemedicine. They will be less likely to accept the technology if they cannot comprehend its potential benefits.

The usage of telemedicine was also shown to be influenced by language distinctions and cultural factors. Practitioners reported that due to the diversity of languages and cultural traits in India, virtual assessment, in general, may become more complex and predisposed to erroneous judgments ^44,63^.

Age was also another socio-cultural factor. Some studies have reported that elderly patients are wary of new technologies and are uncomfortable dealing with computers and other contemporary technologies ^16,26^.

#### Facilitators

Many articles with video teleconferencing and mobile consultation interventions reported telemedicine as safe, acceptable, and effective for consultations in tertiary care ^26,27,37,51,52,54,55,62,64^ despite preconceived doubts about the efficacy of telemedicine consultations. Another important facilitator reported at this level is that telemedicine consultations helped reduce the caregiver burden ^30,36,56^.

The review findings also suggested that stakeholders’ families, neighbours, and peers influence the intention to adopt telemedicine. A study ^63^ found that social influence, through widespread adoption and use of a telemedicine intervention (99DOTS) by colleagues and supervisors, boosted communication among healthcare practitioners and improved their acceptance of the intervention.

### 4.3 egulatory and financial factors

The regulatory level entails the protocols, procedures and functions that enable a telemedicine system to deliver high-quality healthcare services to the community efficiently. This review reports the regulatory and financial barriers and facilitators from all perspectives.

#### Barriers

Data privacy and security risks associated with telemedicine are the most widely reported barriers and are reported as significant barriers to the delivery of primary and secondary care ^26,30,31,43,46,48,51,52,56,58,61^ in all modes of real-time delivery. The adoption of telemedicine in India was reported to be impacted by the security, safety, and confidentiality of patient data ^11,30,31,43,46,48,51,52,55-58^. In telemedicine systems, some patients expressed unwillingness to have their pictures and videos taken due to discomfort with having their images taken, or reluctance to share them with other healthcare providers or worry that their information might be misplaced, stolen, or leaked on social media, or viewed by unauthorised individuals ^52^. Most studies also indicate that doctors are still unsure whether telemedicine is a secure method to communicate sensitive patient data and records and are concerned that unauthorised parties could access the medical data. Additionally, doctors also fear legal issues from the unintentional sharing of patient data. Thus, another significant barrier reported by the stakeholders is the fear of legal liability from malpractice due to the lack of detailed information concerning malpractice liability in the telemedicine guidelines ^11,29-31,48,57,60,62^. The articles dealing with teleconsultations and video teleconferencing interventions indicate these ambiguities concerning the regulations in the Indian telemedicine framework and the consequent legal liabilities as critical barriers.

In addition to the regulatory factors, financial factors also play a crucial role in telemedicine adoption. Every component of the health system, including service delivery, human resources, technology, products, and regulation, is supported by financing. Consequently, the financial impact of telemedicine is influenced by various infrastructural, social, cultural, regulatory, and economic factors. In this review, 35% of the included studies reported that telemedicine systems are perceived to be a financial burden to healthcare providers and patients from all perspectives ^29,30,36,50,57,58,64^. The cost of purchasing and maintaining equipment, training healthcare workers, and paying for recurrent expenses like internet and electricity bills are cited as barriers to implementing telemedicine services. It is perceived that providers typically require financial backing from the government or development partners to make telemedicine operations sustainable, and without adequate support, telemedicine initiatives may not succeed due to the high costs associated with purchasing, installing, and maintaining the technology ^36,60^. Some socio-cultural and infrastructural barriers like limited telemedicine awareness among the patients and local healthcare professionals, low ICT literacy, and restricted access to infrastructure and technology are reported to be the reasons for the perception that telemedicine is costly ^11,22,26,29,30,36,50,58-61,64^.

Furthermore, some studies have also reported the lack of business and/or financial models to integrate telemedicine into the existing healthcare system as a critical barrier to adopting telemedicine systems at this level ^49,58,64,67^. Nevertheless, this study ^43^ reported another interesting finding from the survey of government and public hospital doctors in North India about doctors’ belief that other risks, such as privacy and security risk, social risk, and technological risk, far outweigh the financial risk. The respondents in the study indicated that the perception of financial risk would not prevent their adoption of telemedicine compared to the other risks.

#### Facilitators

The cost of telemedicine systems is reported as one of the significant facilitators and barriers from all perspectives. 59% of the articles reported reduction in healthcare costs as one of the significant facilitators of telemedicine adoption, especially in primary and tertiary healthcare systems, as telemedicine saves time, travel, and the related costs when an in-person consultation is not warranted ^11,16,26,27,30,31,46,49,51-58,61,62,64^.

Another important facilitator stated by the doctors and patients at this level is the extensive increase in the accessibility of quality healthcare to patients through telemedicine. This enhanced patient access to healthcare is reported as another critical facilitator to the adoption of telemedicine in primary ^26,31,43,44,49,50,56-58^ and tertiary healthcare tiers ^27,29,55,64^ delivered through mobile consultation and video teleconferencing interventions. Access to specialist care is one of the key facilitators to adopting telemedicine since it enhances patients’ access to healthcare specialists despite a dearth of specialists in some rural and remote regions. The respondents in this study ^46^ largely agreed that telemedicine would assist in delivering quality healthcare to rural and remote areas of the country. Telemedicine is perceived to be a helpful mode of healthcare delivery in all three healthcare tiers, where equitable healthcare services remain a challenge ^22,26,29-31,49,57,58,64^.

Additionally, the safety of the patients and healthcare providers during a pandemic and the consequent extensive outreach of the hospital due to telemedicine programmes is a facilitating factor from the viewpoints of healthcare organisations ^30^.

Grouping the barriers and facilitators into multiple categories can help understand the factors influencing telemedicine adoption. Table IV presents a synthesis of the key barriers and facilitators, and Table V categorises barriers and facilitators by study perspective, healthcare tier, and mode of delivery.

**Table IV:** Summary of review outcomes.

|  |  |  |
| --- | --- | --- |
| **Infrastructural factors** | **Key Facilitators** | 1. Ease of consultation using telemedicine systems (***all healthcare tiers***)<br>2. Reduced commute to hospitals for consultations (***all healthcare tiers***) |
|  |  | 3. Reduction in travel time and effort from telemedicine appointments (*secondary and tertiary tiers*) |
|  | **Key Barriers** | Lack of reliable internet connectivity and hardware infrastructure (*primary healthcare, all modes of real-time delivery*) |
| **Socio-cultural factors** | **Key Facilitators** | Telemedicine was acceptable and effective for consultations (*tertiary healthcare*) |
|  | **Key Barriers** | 1. Patients' lack of trust in the reliability of the virtual assessment (*secondary healthcare*)<br>2. Preconceived doubts regarding the quality and efficacy of virtual consultations (*video teleconferencing and mobile consultations*) |
| **Regulatory and financial factors** | **Key Facilitators** | 1. Reduction in healthcare delivery costs from telemedicine (*all stakeholder perspectives, primary and tertiary tiers, all modes of real-time delivery*)<br>2. Enhanced access to healthcare for patients in remote areas (*all healthcare tiers*) |
|  | **Key Barriers** | 1. Telemedicine is reported as a financial burden (*all stakeholder perspectives*)<br>2. Fear of violation of the privacy and security of the patient's medical data (*primary and secondary tiers*) |

**Table V:** Facilitators and barriers for telemedicine systems observed from different perspectives at various levels, healthcare tiers, and modes of delivery

| Factors | Facilitators | Doctors & Healthcare Providers | Patients & Patient Caregivers | Society | Healthcare Organisations | Government |
| --- | --- | --- | --- | --- | --- | --- |
| Infrastructural | Time-saving | ✓(T),(VTC) | ✓(H),(VTC,MC) | ✓(P,T),(C) |  |  |
|  | Less commute | ✓(S,T),(VTC) | ✓(H),(VTC,MC) | ✓(P,T),(C) |  |  |
|  | Reduced hospital visits for patients | ✓(H),(C,VTC,MC) | ✓(H),(D) | ✓(H),(C) |  |  |
|  | Ease of consultation for doctors/patients | ✓(H),(C,VTC,MC) | ✓(H),(C,MC) |  |  |  |
|  | Access to high-speed internet and other technology at the healthcare facilities | ✓(P) |  |  |  |  |
|  | Requires only minimal equipment/ User-friendly technology |  | ✓(P,S),(MC) |  |  |  |
|  | Reduced waiting times for the patient | ✓(P,S),(C) | ✓(S),(C,VTC) |  |  |  |
| Socio-Cultural | Lesser chances of contracting hospital-acquired infections | ✓(P),(MC) | ✓(P,S),(C,VTC,MC) |  |  |  |
|  | Convenient for differently abled |  | ✓(S),(VTC) | ✓(P) |  |  |
|  | Financial risk did not affect the adoption | ✓(P),(C) | ✓(T),(MC) |  |  |  |
|  | Technology issues and language barriers did not affect the adoption of telemedicine |  | ✓(T),(MC) |  |  |  |
|  | Safe, acceptable, and effective | ✓(S,T),(VTC,MC) | ✓(H),(VTC,MC) | ✓(P,T),(C) |  |  |
|  | Higher patient satisfaction |  | ✓(S),(VTC) |  |  |  |
|  | Improved patient's mental and physical quality of life |  | ✓(S,T),(VTC,MC) |  |  |  |
|  | Physicians' habit of providing in-person consultations does not affect the adoption of telemedicine | ✓(P),(C) |  |  |  |  |
|  | Patients' habit of seeking in-person consultations does not affect the adoption of telemedicine |  | ✓(P),(C) |  |  |  |
|  | Less caregiver burden |  | ✓(P,S),(C,VTC,MC) | ✓(P,T),(C) |  |  |
|  | Reduced stigma in certain clinical specialties |  | ✓(P) | ✓(P),(C) |  |  |
| **Regulatory & Financial** | Reduced healthcare costs | ✓(H),(C,VTC,MC) | ✓(H),(C,VTC,MC) | ✓(P,T),(C) | ✓(P) | ✓(P) |
|  | Reduced possibility of medical data omission | ✓(P) |  |  |  |  |
|  | Enhanced quality of care and effectiveness | ✓(S),(NC) | ✓(S),(VTC) |  | ✓(P) |  |
|  | Improved patient-provider relationship | ✓(S),(NC) |  | ✓(P) |  |  |
|  | Improved patient access to quality healthcare in remote areas | ✓(H),(C,VTC,MC) | ✓(H),(VTC,MC) | ✓(H) |  | ✓(P) |
|  | Improved patient access to healthcare (especially follow-up) | ✓(H),(C,VTC,MC) | ✓(H),(C,VTC,MC) | ✓(P,T),(C) |  |  |
|  | Simultaneous and easy delivery of information and instructions to several caregivers | ✓(P),(C) |  |  |  |  |
|  | Enhanced safety for patients and healthcare providers |  |  |  | ✓(P) |  |
|  | Increased reach to hospital |  |  |  | ✓(P) |  |
| **Factors** | **Barriers** | **Doctors & Healthcare Providers** | **Patients & Patient Caregivers** | **Society** | **Healthcare Organisations** | **Government** |
| **Infrastructural** | Time-consuming | ✗(P,S),(MC,NC) | ✗(P,S),(MC) |  |  |  |
|  | Unavailability of technology resources | ✖(P,S),(C,MC) | ✖(P,S),(D) | ✖(P,T),(C,VTC) |  | ✖(P) |
|  | Inadequate internet connectivity | ✖(P,S),(MC) | ✖(H),(VTC,MC) | ✖(H),(VTC) |  | ✖(P) |
|  | Technology issues (system installation and maintenance issues) | ✖(P,S),(C,MC) | ✖(P,S),(C,VTC) | ✖(P,T),(VTC) |  |  |
|  | Lack of awareness and expertise among health professionals | ✖(P,S),(C,VTC,MC) | ✖(S),(MC) | ✖(P) |  |  |
|  | Lack of skilled professionals | ✖(P,S),(MC) | ✖(P,S),(C,VTC) | ✖(P,T),(C,VTC) | ✖(P) |  |
|  | Lack of availability and expertise in telemedicine interventions |  | ✖(S),(MC) | ✖(P) |  |  |
| **Socio-Cultural** | Need for physical examination and closer observation | ✖(P,S),(C,VTC) | ✖(P,S),(C,VTC,MC) | ✖(P,T),(C) |  |  |
|  | Poor health/technology literacy (in remote and rural areas) | ✖(P,S),(C,VTC,MC) | ✖(H),(VTC,MC) | ✖(H),(C) |  | ✖(P) |
|  | Patient socioeconomic factors | ✖(S) |  |  |  |  |
|  | Demotivated patients | ✖(S),(MC) |  |  |  |  |
|  | Cultural and language barriers | ✖(S),(MC) | ✖(H),(VTC,MC) | ✖(P,T),(C,VTC) |  | ✖(P) |
|  | Need for social acceptance of telemedicine | ✖(P),(C) | ✖(P,T),(C,VTC) |  |  |  |
|  | Patients' doubts about virtual assessment quality and acceptance | ✖(S),(MC) | ✖(H),(C,VTC,MC) | ✖(H),(C,VTC) |  |  |
|  | Healthcare providers' beliefs about efficacy of telemedicine: e.g., not an adequate substitute for in-person evaluations after the pandemic | ✖(P,S),(C,VTC,MC) | ✖(S,T),(VTC,MC) |  |  |  |
|  | Resistance to change | ✖(P),(MC) | ✖(P),(MC) | ✖(P) | ✖(P) |  |
|  | Doubts regarding ease of e-consultations |  | ✖(P),(C) |  |  |  |
|  | Less family support for telemedicine interventions |  | ✕(S),(MC) |  |  |  |
| **Regulatory & Financial** | Lack of solutions to integrate telemedicine into existing healthcare system despite overcrowded outpatient services | ✕(P,T),(VTC) |  | ✕(P,T),(C) |  |  |
|  | Patient 'no shows' | ✕(T),(VTC) |  |  |  |  |
|  | Financial burden | ✕(P,S),(C,VTC,MC) | ✕(P,S),(C,VTC,MC) | ✕(H),(C,VTC) | ✕(P) | ✕(P) |
|  | Fear of violation of data privacy and security | ✕(P,S),(MC) | ✕(H),(C,VTC,MC) | ✕(H),(C) |  |  |
|  | Practitioner credibility and licensing issues |  |  | ✕(P) |  |  |
|  | Legal liability for healthcare providers | ✕(P,S) |  | ✕(P),(C) |  |  |
|  | Uncertainty concerning telemedicine guidelines and their legal ramifications | ✕(H),(C,VTC,MC) | ✕(P,S),(C,VTC) | ✕(H),(C) |  | ✕(P) (Legal complications) |
|  | Lack of external reward (government initiatives and schemes) | ✕(S),(C,MC) |  |  |  |  |

(x) - healthcare tier(s)
(x),(y) – (healthcare tiers),(modes of delivery)
P-Primary, S-Secondary, T-Tertiary, H-All three healthcare tiers
C- Teleconsultation, VTC-Video Teleconferencing, MC-Mobile Teleconsultation, NC- Non-smartphone-based Technology, D-All four delivery modes

A summary of all barriers and facilitators categorised for the three healthcare sectors and delivery modes is included in the supplementary file1.

### 4.5 Key insights and future research directions

In this review, it could not be established whether certain factors were facilitators or barriers. For instance, it could be rationalised that patients and doctors who reported telemedicine as acceptable and effective may still prefer in-person services, in which case acceptability alone may not facilitate telemedicine adoption. Thus, these factors cannot be considered facilitators or barriers to adoption until practitioner and patient perspectives on how individual factors impact telemedicine adoption are researched.

Another significant issue concerning financial factors is that, since most of the included articles are studies/pragmatic viewpoints of telemedicine barriers and facilitators, the selected studies do not elaborate on the direct and indirect medical and non-medical costs and benefits of telemedicine systems in government and private medical institutions. Consequently, there is no clarity as to what is making telemedicine less affordable or if telemedicine is reducing healthcare costs for patients and providers. These undisclosed costs and benefits might lead to the perception that telemedicine is expensive even when it is not. However, economic assessments of telemedicine systems are outside the scope of this review. Therefore, we could not arrive at a fair characterisation of the conditions under which the cost of telemedicine is a barrier or facilitator, as the reviewed articles do not provide a sufficiently detailed delineation of the costs and benefits of incorporating telemedicine into clinical practice.

Further, some studies in the review outlined their viewpoints on the barriers and facilitators to telemedicine among healthcare professionals without mentioning an existing telemedicine intervention in their study settings. Such studies make it difficult to ascertain the experiential or empirical basis for their viewpoints. Also, most of the studies in this review did not include the opinions of healthcare administrators or clinical managers of telemedicine programs whose responses might have altered the current review findings.

There are also no established classifications of barriers and facilitators or nomenclatures for modes of telemedicine delivery. Therefore, it is difficult to organise the results of the selected studies to prioritise commonly occurring barriers and facilitators in different telemedicine systems. Furthermore, the existing research has not answered a lot of essential questions regarding the telemedicine systems which would impact its adoption, such as whether a combination of in-person and telemedicine consultations would be most effective for patient care or the type of patient care services (e.g., preliminary diagnosis, treatment planning, medication review, etc.) that would be benefited from the implementation of telemedicine systems. Determining the answers to these and other such unanswered questions could significantly impact the adoption of telemedicine systems by the stakeholders.

Therefore, future work can focus on conducting studies with a substantial sample of telemedicine stakeholders to obtain their unbiased, comprehensive opinions about the various facets of real-world telemedicine systems discussed above.

## 5. Limitations

Although this review contributes to the current literature by emphasising recent facilitators and barriers for telemedicine systems in India from different perspectives and contexts, it has limitations. This review is predisposed to bias due to the limited number of relevant published studies and has restrictions arising from the chosen studies’ heterogeneity. The various types of publications considered in this review, such as expert opinions and narrative reviews that primarily provide descriptive data or viewpoints lack established methodologies for assessing bias. Thus, we selected the articles after verifying that a reference or clear rationale supported the results. Since human evaluations may have induced bias when publications were evaluated for selection criteria, we addressed this selection bias objectively by having two reviewers (AV and NF) independently analyse each publication, followed by consensus meetings to ensure that only articles with relevant attributes were included.

Another potential drawback of this review is that the COVID-19-related psychosocial problems may have influenced the opinions in studies published during the COVID-19 lockdown. However, since we conducted an article search from 2016, 20% (n=7) of the selected studies were based on telemedicine interventions implemented before COVID-19.

Furthermore, since we obtained heterogeneous qualitative outcomes from the studies, we could not conduct a meta-analysis, and the rationales behind the reported barriers and facilitators could not be compared. We performed only a descriptive and interpretative analysis of the data. Thus, further research can be carried out to quantitatively substantiate the outcomes of this review by analysing the facilitators and barriers through data collection and surveys of the stakeholders’ experiences in telemedicine in hospital settings.

## 6. Conclusion

Telemedicine can potentially have a significant and long-term impact on India’s healthcare systems. Despite the ostensible benefits of telemedicine applications (e.g., reduced healthcare expenditures and improved patient accessibility), telemedicine remains underdeveloped. Because it is widely acknowledged that telemedicine-based programmes have enormous potential to enhance existing healthcare settings in developing countries, this study aims to fill a vital literature gap by identifying and categorising barriers to telemedicine that must be resolved to ensure successful implementation by utilising the broad range of recognised facilitators. Patients and their caretakers, RMPs, nurses, and healthcare and technology providers are the major stakeholders of this system, and their awareness and perceptions of telemedicine might be the key to adopting telemedicine as a fundamental healthcare option. This review will help inform telemedicine stakeholders and will assist relevant policymakers, regulators, and other researchers in identifying gaps in the Indian research and policy landscape around telemedicine that need to be addressed as part of evolving an effective framework for the successful implementation and evaluation of telemedicine systems in the Indian context. These review outcomes may also apply to other developing countries with healthcare delivery landscapes similar to India.

## Data Availability

This work only reviewed and analysed secondary data from the published articles, which are included in this manuscript and the supplementary files.

## Author Contributions

Authors AV, VR, and SE assessed the review’s feasibility and helped define the research question. AV devised the review methodology and the search technique and conducted the search. AV and NF were involved in the study selection and the data extraction process. AV wrote the article, and all authors contributed to the manuscript’s revision.

## References

1. Mathur P, Srivastava S, Lalchandani A, Mehta JL. Evolving Role of Telemedicine in Health Care Delivery in India. Primary Health Care 2017; 7 : 260.

2. Closed database: National Data & Analytics Platform. Population Projections. Jan 2022. Available from: https://ndap-beta.niti.gov.in/dataset/7208

3. Closed database: National Data & Analytics Platform. Population Projections Urban. Jan 2022. Available from: https://ndap-beta.niti.gov.in/dataset/7209

4. Closed database: National Data & Analytics Platform. Handbook of Statistics on Indian States: Infrastructure Doctors and Specialists. Jan 2022. Available from: https://ndap-beta.niti.gov.in/dataset/7053

5. WHO Global Observatory for eHealth. Telemedicine: opportunities and developments in Member States: report on the second global survey on eHealth. Geneva: World Health Organization; 2010.

6. De La Torre-Díez I, López-Coronado M, Vaca C, Aguado JS, De Castro C. Cost-Utility and Cost-Effectiveness Studies of Telemedicine, Electronic, and Mobile Health Systems in the Literature: A Systematic Review. Telemedicine and e-Health 2015; 21 (2) : 81–85.

7. Kaeley N, Choudhary S, Mahala P, Nagasubramanyam V. Current scenario, future possibilities and applicability of telemedicine in hilly and remote areas in India: A review protocol. J Family Med Prim Care 2021; 10 (1) : 77–83.

8. Ministry of External Affairs GoI [homepage on the Internet]. Available from: http://www.mea.gov.in/, accessed on April 15, 2023.

9. Agarwal N, Jain P, Pathak R, Gupta R. Telemedicine in India: A tool for transforming health care in the era of COVID-19 pandemic. Journal of education and health promotion 2020; 9 : 190

10. Board of Governors-Indian Medical Council. Telemedicine Practice Guidelines Enabling Registered Medical Practitioners to Provide Healthcare Using Telemedicine. Indian Medical Council 2020.

11. Mahajan V, Singh T, Azad C. Using Telemedicine During the COVID-19 Pandemic. Indian Pediatrics 2020; 57 (7) : 658–661.

12. Agarwal N, Biswas B. Doctor Consultation through Mobile Applications in India: An Overview, Challenges and the Way Forward. Healthc Inform Res 2020; 26 (2) : 153–158.

13. Dash S, Aarthy R, Mohan V. Telemedicine during COVID-19 in India—a new policy and its challenges. Journal of Public Health Policy: Palgrave Macmillan UK; 2021.

14. Garg S, Gangadharan N, Bhatnagar N, Singh MM, Raina SK, Galwankar S. Telemedicine: Embracing virtual care during COVID-19 pandemic. Journal of family medicine and primary care 2020; 9 : 4516–4520.

15. Dinakaran D, Manjunatha N, Kumar CN, Math SB. Telemedicine practice guidelines of India, 2020: Implications and challenges. Indian journal of psychiatry 2020; 63 : 97–101.

16. Appireddy R, Bendahan N, Chaitanya J, Shukla G. Virtual Care for Neurological Practice. Ann Indian Acad Neurol 2020; 23 (5) : 587–591.

17. Chandrasekaran S, Chandrashekar VS, Dalvie S, Sinha A. The case for the use of telehealth for abortion in India. Sex Reprod Health Matters 2021; 29 (2) : 1920566.

18. Deshpande S, Patil D, Dhokar A, Bhanushali P, Katge F. Teledentistry: A Boon Amidst COVID-19 Lockdown-A Narrative Review. Int J Telemed Appl 2021; 2021 : 8859746.

19. Galagali PM, Ghosh S, Bhargav H. The Role of Telemedicine in Child and Adolescent Healthcare in India. Curr Pediatr Rep 2021; 9 (4) : 1–8.

20. Gupta R, Kumar VM, Tripathi M, Datta K, Narayana M, Ranjan Sarmah K, et al. Guidelines of the Indian Society for Sleep Research (ISSR) for Practice of Sleep Medicine during COVID-19. Sleep Vigil 2020; 4 (2) : 1–12.

21. Jayadev C, Mahendradas P, Vinekar A, Kemmanu V, Gupta R, Pradhan ZS, et al. Teleconsultations in the wake of COVID-19 - Suggested guidelines for clinical ophthalmology. Indian J Ophthalmol 2020; 68 (7) : 1316–1327.

22. Sharma M, Jain N, Ranganathan S, Sharma N, Honavar SG, Sharma N, et al. Tele-ophthalmology: Need of the hour. Indian J Ophthalmol 2020; 68 (7) : 1328–1338.

23. Pasquali P, Sonthalia S, Moreno-Ramirez D, Sharma P, Agrawal M, Gupta S, et al. Teledermatology and its Current Perspective. Indian Dermatol Online J 2020; 11 (1) : 12–20.

24. Adhikari SD, Biswas S, Mishra S, Kumar V, Bharti SJ, Gupta N, et al. Telemedicine as an Acceptable Model of Care in Advanced stage Cancer Patients in the Era of Coronavirus Disease 2019 - An Observational Study in a Tertiary Care Centre. Indian J Palliat Care 2021; 27 (2) : 306–312.

25. Dinakaran D, Basavarajappa C, Manjunatha N, Kumar CN, Math SB. Telemedicine Practice Guidelines and Telepsychiatry Operational Guidelines, India—A Commentary. Indian Journal of Psychological Medicine 2020; 42 (Suppl) : 1S–3S.

26. Vadlamani LN, Sharma V, Emani A, Gowda MR. Telepsychiatry and Outpatient Department Services. Indian J Psychol Med 2020; 42 (Suppl) : 27S–33S.

27. Jayarajan D, Sivakumar T, Torous JB, Thirthalli J. Telerehabilitation in Psychiatry. Indian J Psychol Med 2020; 42 (Suppl) : 57S–62S.

28. Pradeepa R, Rajalakshmi R, Mohan V. Use of Telemedicine Technologies in Diabetes Prevention and Control in Resource-Constrained Settings: Lessons Learned from Emerging Economies. Diabetes Technol Ther 2019; 21 (Suppl 2) : S2–9-16.

29. Ramakrishnan N, Tirupakuzhi Vijayaraghavan BK, Venkataraman R. Breaking Barriers to Reach Farther: A Call for Urgent Action on Tele-ICU Services. Indian J Crit Care Med 2020; 24 (6) : 393–397.

30. Sharma RK, Prashar R. Feasibility of eHealth Implementation in India Learning from Global Experience. Asia Pacific Journal of Health Management 2019; 14 (3) : 12–23.

31. Verma V, Krishnan V, Verma C. Telemedicine in India - an investment of technology for a digitized healthcare industry: a systematic review. Revista Română de Informatică şi Automatică 2021; 31 (4) : 33–44.

32. Chandwani RK, Dwivedi YK. Telemedicine in India: current state, challenges and opportunities. Transforming Government: People, Process and Policy 2015; 9 (4) : 393–400.

33. Aparna V, Ramamohan V, Edirippulige S. Analysis of modern barriers and facilitators for telemedicine systems in India. PROSPERO; 2022.

34. Davies KS. Formulating the Evidence Based Practice Question: A Review of the Frameworks. Evidence Based Library and Information Practice 2011; 6 (2) : 75–80.

35. Page MJ, Moher D, Bossuyt PM, Boutron I, Hoffmann TC, Mulrow CD, et al. PRISMA 2020 explanation and elaboration: updated guidance and exemplars for reporting systematic reviews. BMJ 2021; 372 : 160.

36. Bairapareddy KC, Alaparthi GK, Jitendra RS, Prathiksha Rao PP, Shetty V, et al. “We are so close; yet too far”: perceived barriers to smartphone-based telerehabilitation among healthcare providers and patients with Chronic Obstructive Pulmonary Disease in India. Heliyon 2021; 7 (8) : e07857.

37. Hameed BZ, Shah M, Naik N, Reddy SJ, Somani BK. Use of ureteric stent related mobile phone application (UROSTENTZ App) in COVID-19 for improving patient communication and safety: a prospective pilot study from a university hospital. Cent European J Urol 2021; 74 (1) : 51–56.

38. Barigela R, Kodali PB, Hense S. What is Stopping Primary Health Centers to Go Digital? Findings of a Mixed-method Study at a District Level Health System in Southern India. Indian J Community Med 2021; 46 (1) : 97–101.

39. Gopalakrishnan L, Buback L, Fernald L, Walker D, Diamond-Smith N, The CAS Evaluation Consortium. Using mHealth to improve health care delivery in India: A qualitative examination of the perspectives of community health workers and beneficiaries. PLoS ONE 2020; 15 (1) : e0227451.

40. Andrees V, Klein TM, Augustin M, Otten M. Live interactive teledermatology compared to in-person care - a systematic review. J Eur Acad Dermatol Venereol 2020; 34 (4) : 733–745.

41. Ajay VS, Jindal D, Roy A, Venugopal V, Sharma R, Pawar A, et al. Development of a Smartphone-Enabled Hypertension and Diabetes Mellitus Management Package to Facilitate Evidence-Based Care Delivery in Primary Healthcare Facilities in India: The mPower Heart Project. J Am Heart Assoc 2016; 5 (12).

42. Wani RT. Socioeconomic status scales-modified Kuppuswamy and Udai Pareekh’s scale updated for 2019. J Family Med Prim Care 2019; 8 (6) : 1846–1849.

43. Bakshi S, Tandon U. Understanding barriers of telemedicine adoption: A study in North India. Systems Research and Behavioral Science 2021; 39 (1) : 128–142.

44. Banerjee D, Vajawat B, Varshney P, Rao TS. Perceptions, Experiences, and Challenges of Physicians Involved in Dementia Care During the COVID-19 Lockdown in India: A Qualitative Study. Front Psychiatry 2020; 11 : 615758.

45. Corley J. Telestroke: India’s solution to a public health-care crisis. Lancet Neurol 2018; 17 (2) : 115–116.

46. Das N, Narnoli S, Kaur A, Sarkar S, Balhara YPS. Attitude to telemedicine in the times of COVID-19 pandemic: Opinion of medical practitioners from India. Psychiatry Clin Neurosci 2020; 74 (10) : 560–562.

47. Dash A, Sahoo AK. Physician’s perception of E-consultation adoption amid of COVID-19 pandemic. VINE Journal of Information and Knowledge Management Systems 2021; (ahead-of-print).

48. Iyengar K, Jain VK, Vaishya R. Pitfalls in telemedicine consultations in the era of COVID 19 and how to avoid them. Diabetes & Metabolic Syndrome: Clinical Research & Reviews 2020; 14 (5) : 797–799.

49. Joshi NK, Bhardwaj P, Suthar P, Jain YK, Joshi V, Singh K. Overview of e-Health initiatives in Rajasthan: An exploratory study. J Family Med Prim Care 2021; 10 (3) : 1369–1376.

50. Dash A, Sahoo AK. Moderating effect of gender on adoption of digital health consultation: a patient perspective study. International Journal of Pharmaceutical and Healthcare Marketing 2021; 15 (4) : 598–616.

51. Kanuri N, Arora P, Talluru S, Colaco B, Dutta R, Rawat A, et al. Examining the initial usability, acceptability and feasibility of a digital mental health intervention for college students in India. Int J Psychol 2020; 55 (4) : 657–673.

52. Kumar AA, De Costa A, Das A, Srinivasa GA, D’Souza G, Rodrigues R. Mobile Health for Tuberculosis Management in South India: Is Video-Based Directly Observed Treatment an Acceptable Alternative? JMIR Mhealth Uhealth 2019; 7 (4) : e11687.

53. Verma N, Mishra S, Singh S, Kaur R, Kaur T, De A, et al. Feasibility, Outcomes, and Safety of Telehepatology Services During the COVID-19 Pandemic. Hepatol Commun 2022; 6 (1) : 65–76.

54. Manglani M, Gabhale Y, Lala MM, Balakrishnan S, Bhuyan K, Rewari BB, et al. Reaching the Unreached: Providing Quality Care to HIV-Infected Children through Telemedicine—An Innovative Pilot Initiative from Maharashtra, India. International Journal of Pediatrics 2020; 2020 : 1–11.

55. Mahmood A, Blaizy V, Verma A, Stephen Sequeira J, Saha D, Ramachandran S, et al. Acceptability and Attitude towards a Mobile-Based Home Exercise Program among Stroke Survivors and Caregivers: A Cross-Sectional Study. Int J Telemed Appl 2019; 2019 : 5903106.

56. Naik SS, Manjunatha N, Kumar CN, Math SB, Moirangthem S. Patient’s Perspectives of Telepsychiatry: The Past, Present and Future. Indian J Psychol Med 2020; 42 (Suppl) : 102S–107S.

57. Ateriya N, Saraf A, Meshram VP, Setia P. Telemedicine and virtual consultation: The Indian perspective. Natl Med J India 2018; 31 (4) : 215–218.

58. Bali S. Barriers to Development of Telemedicine in Developing Countries. In: Heston TF, editor. Telehealth. IntechOpen; 2019. p. 1–14.

59. Biswas P, Batra S. Commentary: Telemedicine: The unsung corona warrior. Indian Journal of Ophthalmology 2020; 68 (6).

60. Ghai S. Teledentistry during COVID-19 pandemic. Diabetes Metab Syndr 2020; 14 (5) : 933–935.

61. Kalaivanan RC, Rahul P, Manjunatha N, Kumar CN, Sivakumar PT, Math SB. Telemedicine in Geriatric Psychiatry: Relevance in India. Indian J Psychol Med 2021; 43 (5S): 121S–127S.

62. Kavadichanda C, Shah S, Daber A, Bairwa D, Mathew A, Dunga S, et al. Tele-rheumatology for overcoming socioeconomic barriers to healthcare in resource constrained settings: lessons from COVID-19 pandemic. Rheumatology (Oxford, England) 2021; 60 (7) : 3369–3379.

63. Thomas BE, Kumar JV, Onongaya C, Bhatt SN, Galivanche A, Periyasamy M, et al. Explaining Differences in the Acceptability of 99DOTS, a Cell Phone-Based Strategy for Monitoring Adherence to Tuberculosis Medications: Qualitative Study of Patients and Health Care Providers. JMIR Mhealth Uhealth 2020; 8 (7) : e16634.

64. Chandwani R, Kumar N. Stitching Infrastructures to Facilitate Telemedicine for Low-Resource Environments. Proceedings of the 2018 CHI Conference on Human Factors in Computing Systems; 2018 Apr 21–26; Montréal, QC, Canada. ACM; 2018.

65. Mittal A, Pareek P. Telephonic Triage and Telemedicine During the Peak of COVID-19 Pandemic — Restricting Exposure to Healthcare Professionals. Indian Pediatrics 2020; 57 (10) : 973–974.

66. Bali S, Gupta A, Khan A, Pakhare A. Evaluation of telemedicine centres in Madhya Pradesh, Central India. Journal of Telemedicine and Telecare 2016; 22 (3) : 183–188.

67. Singh Pardal MP, Rajiva K, Orkeh GO. Telemedicine in the era of COVID-19: The East and the West. J Mar Med Soc 2020; 22 : S32–5.

