## Supplementary File1 for "Facilitators and Barriers for Telemedicine Systems in India from Multiple Stakeholder Perspectives and Settings"

**Keyword groups for the search query**

| **Code** | **Group** | **Keywords** |
| --- | --- | --- |
| 1 | Barriers and Facilitators | barrier(s), obstacle(s), hindrance(s), hinder, obstructions(s), obstructors, impediment(s), facilitator(s) |
| 2 | Primary Healthcare | primary healthcare, primary care, general physician, general practice, family practice, family medicine |
| 3 | Secondary Healthcare | secondary care, specialist(s), specialist care |
| 4 | Tertiary Healthcare | tertiary healthcare, tertiary care, Surgery, Tertiary Hospital, Tertiary Referral centre, Emergency care |
| 5 | Telemedicine | Telemedicine, Telehealth, remote consultation, teleconsultation, Telemonitoring, Telecare, Telehealthcare, Telehomecare, ehealth, Uhealth, Mhealth, digital health, econsultation |
| 6 | India | India |

**Search results from the databases for the review period**

| **Database** | **Keyword Group Codes** | **Refinements** | **Results** |
| --- | --- | --- | --- |
| PubMed | 1 AND 5 AND 6 | 1. Articles published during the last five years  2. Articles with full-text availability  3. Articles dealing with human population | 148 |
| Cochrane | 1 AND 5 AND 6 | Search with different variations of keywords | 260 |
| Scopus | 1 AND 2 AND 3 AND 4 AND 5 AND 6 | Articles published during the last five years | 466 |
| Web of Science | 1 AND 5 AND 6 | Articles published during the last five years | 355 |
| CINAHL | 1 AND 5 AND 6 | 1. Peer-reviewed articles  2. Articles dealing with human population | 45 |
| MEDLINE | 1 AND 5 AND 6 | 1. Peer-reviewed articles  2. Articles dealing with human population | 475 |
| PsycInfo | 1 AND 5 AND 6 | 1. Articles published during the last five years  2. Peer-reviewed articles  3. Articles with full-text availability | 1 |

**PubMed**

(((((((((barrier) OR (obstacle)) OR (hindrance)) OR (hinder)) OR (obstructions)) OR (obstructors)) OR (impediment)) OR (facilitator))

AND

((((((((((((((((((((((((((Telemedicine) OR (Tele-medicine)) OR (Telehealth)) OR (Tele-health)) OR (remote consultation)) OR (teleconsultation)) OR (tele-consultation)) OR (Telemonitoring)) OR (Tele-monitoring)) OR (Telecare)) OR (Tele-care)) OR (Telehealthcare)) OR (Tele-healthcare)) OR (Tele-health care)) OR (Telehomecare)) OR (Tele-homecare)) OR (Tele-home care)) OR (ehealth)) OR (e-health)) OR (Uhealth)) OR (U-health)) OR (Mhealth)) OR (M-health)) OR (digital health)) OR (e-consultation)) OR (econsultation)))

AND

(India)

**Cochrane**

barrier* OR obstacle* OR hindrance* OR hinder* OR obstruction* OR obstructor* OR impediment* OR facilitator*

AND

[Telemedicine] OR "telemedicine" OR "telemedicine's" OR "Tele-medicine" OR ("telehealth's" OR [Telemedicine] OR "telemedicine" OR "telehealth") OR "Tele-health" OR ([Remote Consultation] OR ("remote" AND "consultation") OR "remote consultation") OR ([Remote Consultation] OR ("remote" AND "consultation") OR "remote consultation" OR "teleconsultation" OR "teleconsultations" OR "teleconsult" OR "teleconsultant" OR "teleconsultants" OR "teleconsultative" OR "teleconsulting" OR "teleconsults") OR "tele-consultation" OR ("telemonitor" OR "telemonitored" OR "telemonitoring" OR "telemonitors") OR "Tele-monitoring" OR "Telecare" OR "Tele-care" OR "Telehealthcare" OR "Tele-healthcare" OR ("Tele-health" AND "care") OR "Telehomecare" OR "Tele-homecare" OR ("Tele-home" AND "care") OR ([Telemedicine] OR "telemedicine" OR "ehealth") OR "e-health" OR "Uhealth" OR "U-health" OR ("mhealth's" OR [Telemedicine] OR "telemedicine" OR "mhealth") OR "M-health" OR (("digital" AND "health") OR "digital health") OR "e-consultation" OR ("econsult" OR "econsultation" OR "econsultations" OR "econsults")

AND

[India] OR "india" OR "india's" OR "indias"

**Scopus**

ALL ( barrier ) OR ALL ( "barrier's" ) OR ALL ( barriers ) OR ALL ( obstacle ) OR ALL ( obstacles ) OR ALL ( hindrance ) OR ALL ( hindrances ) OR ALL ( hinder ) OR ALL ( hindered ) OR ALL ( hindering ) OR ALL ( hinders ) OR ALL ( obstruct ) OR ALL ( obstructed ) OR ALL ( obstructing ) OR ALL ( obstruction ) OR ALL ( obstructions ) OR ALL ( obstructive ) OR ALL ( obstructs ) OR ALL ( obstructors ) OR ALL ( impediment ) OR ALL ( impediments ) OR ALL ( facilitate ) OR ALL ( facilitated ) OR ALL ( facilitates ) OR ALL ( facilitating ) OR ALL ( facilitation ) OR ALL ( facilitations ) OR ALL ( facilitative ) OR ALL ( facilitator ) OR ALL ( "facilitator s" ) OR ALL ( facilitators )

AND

INDEXTERMS ( "primary health care" ) OR ( ALL ( primary ) AND ALL ( health ) AND ALL ( care ) ) OR ALL ( "primary health care" ) OR ( ALL ( primary ) AND ALL ( healthcare ) ) OR ALL ( "primary healthcare" ) OR ( ALL ( primary ) AND ALL ( care ) ) OR ALL ( "primary care" ) OR INDEXTERMS ( "general practitioners" ) OR ( ALL ( general ) AND ALL ( practitioners ) ) OR ALL ( "general practitioners" ) OR ( ALL ( general ) AND ALL ( physician ) ) OR ALL ( "general physician" ) OR INDEXTERMS ( "general practice" ) OR ( ALL ( general ) AND ALL ( practice ) ) OR ALL ( "general practice" ) OR INDEXTERMS ( "family practice" ) OR ( ALL ( family ) AND ALL ( practice ) ) OR ALL ( "family practice" ) OR ( ALL ( family ) AND ALL ( medicine ) ) OR ALL ( "family medicine" )

AND

INDEXTERMS ( "secondary care" ) OR ( ALL ( secondary ) AND ALL ( care ) ) OR ALL ( "secondary care" ) OR ALL ( specialist's ) OR ALL ( specialistic ) OR INDEXTERMS ( specialization ) OR ALL ( specialization ) OR ALL ( specialist ) OR ALL ( specialists ) OR ( ( ALL ( specialist's ) OR ALL ( specialistic ) OR INDEXTERMS ( specialization ) OR ALL ( specialization ) OR ALL ( specialist ) OR ALL ( specialists ) ) AND ALL ( care ) ) OR ALL ( telecardiology ) OR ALL ( tele-cardiology ) OR ALL ( teleobstetrics ) OR ALL ( tele-obstetrics ) OR ALL ( telepsychiatry ) OR ALL ( tele-psychiatry ) OR ALL ( teledermatology ) OR ALL ( teledermatology's ) OR ALL ( tele-dermatology ) OR ALL ( telepediatric ) OR ALL ( tele-pediatric ) OR ALL ( teleophthalmology ) OR ALL ( tele-ophthalmology ) OR ALL ( teleoptometry ) OR ALL ( tele-optometry ) OR ALL ( telephysiotherapy ) OR ALL ( tele-physiotherapy ) OR INDEXTERMS ( telerehabilitation ) OR ALL ( telerehabilitation ) OR ( ALL ( tele ) AND ALL ( rehabilitation ) ) OR ALL ( "tele rehabilitation" ) OR ALL ( teleendocrinology ) OR ALL ( tele-endocrinology ) OR ALL ( teleoncology ) OR ALL ( tele-oncology )

AND

INDEXTERMS ( "tertiary healthcare" ) OR ( ALL ( tertiary ) AND ALL ( healthcare ) ) OR ALL ( "tertiary healthcare" ) OR ( ALL ( tertiary ) AND ALL ( care ) ) OR ALL ( "tertiary care" ) OR INDEXTERMS ( surgery ) OR ALL ( surgery ) OR INDEXTERMS ( "surgical procedures, operative" ) OR ( ALL ( surgical ) AND ALL ( procedures ) AND ALL ( operative ) ) OR ALL ( "operative surgical procedures" ) OR INDEXTERMS ( "general surgery" ) OR ( ALL ( general ) AND ALL ( surgery ) ) OR ALL ( "general surgery" ) OR ALL ( surgery's ) OR ALL ( surgerys ) OR ALL ( surgeries ) OR INDEXTERMS ( "tertiary care centers" ) OR ( ALL ( tertiary ) AND ALL ( care ) AND ALL ( centers ) ) OR ALL ( "tertiary care centers" ) OR ( ALL ( tertiary ) AND ALL ( hospital ) ) OR ALL ( "tertiary hospital" ) OR ( ALL ( tertiary ) AND ALL ( referral ) AND ALL ( centre ) ) OR ALL ( "tertiary referral centre" ) OR INDEXTERMS ( "emergency treatment" ) OR ( ALL ( emergency ) AND ALL ( treatment ) ) OR ALL ( "emergency treatment" ) OR ( ALL ( emergency ) AND ALL ( care ) ) OR ALL ( "emergency care" ) OR INDEXTERMS ( "emergency medical services" ) OR ( ALL ( emergency ) AND ALL ( medical ) AND ALL ( services ) ) OR ALL ( "emergency medical services" )

AND

INDEXTERMS ( telemedicine ) OR ALL ( telemedicine ) OR ALL ( "telemedicine's" ) OR ALL ( tele-medicine ) OR ( ALL ( "telehealth s" ) OR INDEXTERMS ( telemedicine ) OR ALL ( telemedicine ) OR ALL ( telehealth ) ) OR ALL ( tele-health ) OR ( INDEXTERMS ( "remote consultation" ) OR ( ALL ( remote ) AND ALL ( consultation ) ) OR ALL ( "remote consultation" ) ) OR ( INDEXTERMS ( "remote consultation" ) OR ( ALL ( remote ) AND ALL ( consultation ) ) OR ALL ( "remote consultation" ) OR ALL ( teleconsultation ) OR ALL ( teleconsultations ) OR ALL ( teleconsult ) OR ALL ( teleconsultant ) OR ALL ( teleconsultants ) OR ALL ( teleconsultative ) OR ALL ( teleconsulting ) OR ALL ( teleconsults ) ) OR ALL ( tele-consultation ) OR ( ALL ( telemonitor ) OR ALL ( telemonitored ) OR ALL ( telemonitoring ) OR ALL ( telemonitors ) ) OR ALL ( tele-monitoring ) OR ALL ( telecare ) OR ALL ( tele-care ) OR ALL ( telehealthcare ) OR ALL ( tele-healthcare ) OR ( ALL ( tele-health ) AND ALL ( care ) ) OR ALL ( telehomecare ) OR ALL ( tele-homecare ) OR ( ALL ( tele-home ) AND ALL ( care ) ) OR ( INDEXTERMS ( telemedicine ) OR ALL ( telemedicine ) OR ALL ( ehealth ) ) OR ALL ( e-health ) OR ALL ( uhealth ) OR ALL ( u-health ) OR ( ALL ( "mhealth's" ) OR INDEXTERMS ( telemedicine ) OR ALL ( telemedicine ) OR ALL ( mhealth ) ) OR ALL ( m-health ) OR ( SRCTITLE ( "lancet digit health" ) OR SRCTITLE ( "eur heart j digit health" ) OR SRCTITLE ( "digit health" ) OR ( ALL ( digital ) AND ALL ( health ) ) OR ALL ( "digital health" ) ) OR ALL ( e-consultation ) OR ( ALL ( econsult ) OR ALL ( econsultation ) OR ALL ( econsultations ) OR ALL ( econsults ) )

AND

INDEXTERMS ( india ) OR ALL ( india ) OR ALL ( india's ) OR ALL ( indias )

**Web of Science**

ALL=barrier OR ALL="barrier‘s" OR ALL=barriers OR ALL=obstacle OR ALL=obstacles OR ALL=hindrance OR ALL=hindrances OR ALL=hinder OR ALL=hindered OR ALL=hindering OR ALL=hinders OR ALL=obstruct OR ALL=obstructed OR ALL=obstructing OR ALL=obstruction OR ALL=obstructions OR ALL=obstructive OR ALL=obstructs OR ALL=obstructors OR ALL=impediment OR ALL=impediments OR ALL=facilitate OR ALL=facilitated OR ALL=facilitates OR ALL=facilitating OR ALL=facilitation OR ALL=facilitations OR ALL=facilitative OR ALL=facilitator OR ALL="facilitator s" OR ALL=facilitators

AND

"ALL=telemedicine OR ALL=telemedicine OR ALL=""telemedicine s"" OR ALL=Tele-medicine OR (ALL=""telehealth s"" OR ALL=telemedicine OR ALL=telemedicine OR ALL=telehealth) OR ALL=Tele-health OR (ALL=""remote consultation"" OR (ALL=remote AND ALL=consultation) OR ALL=""remote consultation"") OR (ALL=""remote consultation"" OR (ALL=remote AND ALL=consultation) OR ALL=""remote consultation"" OR ALL=teleconsultation OR ALL=teleconsultations OR ALL=teleconsult OR ALL=teleconsultant OR ALL=teleconsultants OR ALL=teleconsultative OR ALL=teleconsulting OR ALL=teleconsults) OR ALL=tele-consultation OR (ALL=telemonitor OR ALL=telemonitored OR ALL=telemonitoring OR ALL=telemonitors) OR ALL=Tele-monitoring OR ALL=Telecare OR ALL=Tele-care OR ALL=Telehealthcare OR ALL=Tele-healthcare OR (ALL=Tele-health AND ALL=care) OR ALL=Telehomecare OR ALL=Tele-homecare OR (ALL=Tele-home AND ALL=care) OR (ALL=telemedicine OR ALL=telemedicine OR ALL=ehealth) OR ALL=e-health OR ALL=Uhealth OR ALL=U-health OR (ALL=""mhealth s"" OR ALL=telemedicine OR ALL=telemedicine OR ALL=mhealth) OR ALL=M-health OR (SO=""lancet digit health"" OR SO=""eur heart j digit health"" OR SO=""digit health"" OR (ALL=digital AND ALL=health) OR ALL=""digital health"") OR ALL=e-consultation OR (ALL=econsult OR ALL=econsultation OR ALL=econsultations OR ALL=econsults)"

AND

ALL=india OR ALL=india OR ALL=india's OR ALL=indias

**CINAHL & MEDLINE**

( barrier OR barrier’s OR barriers OR obstacle OR obstacles OR hindrance OR hindrances OR hinder OR hindered OR hindering OR hinders OR obstruct OR obstructed OR obstructing OR obstruction OR obstructions OR obstructive OR obstructs OR obstructors OR impediment OR impediments OR facilitate OR facilitated OR facilitates OR facilitating OR facilitation OR facilitations OR facilitative OR facilitator OR facilitator’s OR facilitators )

AND

( (MH telemedicine+) OR telemedicine OR telemedicine’s OR Tele-medicine OR (telehealth’s OR (MH telemedicine+) OR telemedicine OR telehealth) OR Tele-health OR ((MH "remote consultation+") OR (remote AND consultation) OR "remote consultation") OR ((MH "remote consultation+") OR (remote AND consultation) OR "remote consultation" OR teleconsultation OR teleconsultations OR teleconsult OR teleconsultant OR teleconsultants OR teleconsultative OR teleconsulting OR teleconsults) OR tele-consultation OR (telemonitor OR telemonitored OR telemonitoring OR telemonitors) OR Tele-monitoring OR Telecare OR Telecare OR Telehealthcare OR Telehealthcare OR (Tele-health AND care) OR Telehomecare OR Tele-homecare OR (Tele-home AND care) OR ((MH telemedicine+) OR telemedicine OR ehealth) OR e-health OR Uhealth OR U-health OR (mhealth’s OR (MH telemedicine+) OR telemedicine OR mhealth) OR M-health OR ((SO "lancet digit health" OR ST "lancet digit health" OR IB "lancet digit health") OR (SO "eur heart j digit health" OR ST "eur heart j digit health" OR IB "eur heart j digit health") OR (SO "digit health" OR ST "digit health" OR IB "digit health") OR (digital AND health) OR "digital health") OR econsultation OR (econsult OR econsultation OR econsultations OR econsults) )

AND

( (MH india+) OR india OR india's OR indias )

**PsycInfo**

(((Any Field: (barrier) OR Any Field: (barriers) OR Any Field: (barriers) OR Any Field: (obstacle) OR Any Field: (obstacles) OR Any Field: (hindrance) OR Any Field: (hindrances) OR Any Field: (hinder) OR Any Field: (hindered) OR Any Field: (hindering) OR Any Field: (hinders) OR Any Field: (obstruct) OR Any Field: (obstructed) OR Any Field: (obstructing) OR Any Field: (obstruction) OR Any Field: (obstructions) OR Any Field: (obstructive) OR Any Field: (obstructs) OR Any Field: (obstructors) OR Any Field: (impediment) OR Any Field: (impediments) OR Any Field: (facilitate) OR Any Field: (facilitated) OR Any Field: (facilitates) OR Any Field: (facilitating) OR Any Field: (facilitation) OR Any Field: (facilitations) OR Any Field: (facilitative) OR Any Field: (facilitator) OR Any Field: (facilitators) OR Any Field: (facilitators)))

AND

(((MeSH: (telemedicine)) OR (Any Field: (telemedicine) OR Any Field: (telemedicine's) OR Any Field: ("Tele-medicine") OR Any Field: (telehealth's) OR Any Field: (telehealth) OR Any Field: ("Tele-health")) OR (MeSH: (remote consultation)) OR (Any Field: (remote) AND Any Field: (consultation)) OR Any Field: ("remote consultation") OR (Any Field: (teleconsultation) OR Any Field: (teleconsultations) OR Any Field: (teleconsult) OR Any Field: (teleconsultant) OR Any Field: (teleconsultants) OR Any Field: (teleconsultative) OR Any Field: (teleconsulting) OR Any Field: (teleconsults) OR Any Field: ("teleconsultation")) OR (Any Field: (telemonitor) OR Any Field: (telemonitored) OR Any Field: (telemonitoring) OR Any Field: (telemonitors) OR Any Field: ("Tele-monitoring") OR Any Field: (Telecare) OR Any Field: ("Tele-care")) OR (Any Field: (Telehealthcare) OR Any Field: ("Tele-healthcare") OR (Any Field: (Tele-health) AND Any Field: (care)) OR Any Field: (Telehomecare) OR Any Field: ("Tele-homecare") OR (Any Field: (Tele-home) AND Any Field: (care))) OR (Any Field: (ehealth) OR Any Field: ("e-health") OR Any Field: (Uhealth) OR Any Field: ("U-health") OR Any Field: (mhealth's) OR Any Field: (mhealth) OR Any Field: ("M-health")) OR (Any Field: (digital) AND Any Field: (health)) OR Any Field: ("digital health") OR Any Field: ("econsultation") OR Any Field: (econsult) OR Any Field: (econsultation) OR Any Field: (econsultations) OR Any Field: (econsults))

AND

(((MeSH: (india)) OR (Any Field: (india) OR Any Field: (india's) OR Any Field: (indias) OR Any Field: (indian)))

#### Summary of the facilitators for telemedicine systems in India

| **Facilitators** | **Count** |
| --- | --- |
| Time-saving | 15 |
| Less commute | 12 |
| Reduced hospital visits for patients | 15 |
| Ease of consultation for doctors/patients | 10 |
| Lesser chances of contracting hospital-acquired infections | 3 |
| Enhanced safety for the patients and healthcare providers | 1 |
| Convenient for differently-abled | 2 |
| Reduced healthcare costs | 20 |
| The financial risk did not affect the adoption of telemedicine | 3 |
| Technology issues and language barriers did not affect the adoption of telemedicine | 1 |
| Access to high-speed internet and other technology at the healthcare facilities | 1 |
| Requirement of minimal equipment / user-friendly technology | 2 |
| Simultaneous and expedited delivery of information and instructions to multiple caregivers | 1 |
| Safe, acceptable, and effective | 14 |
| Higher patient satisfaction | 1 |
| Reduced possibility of medical data omission | 2 |
| Improved efficiency and care quality | 2 |
| Improved patient-provider relationship | 2 |
| Improved patient's mental and physical quality of life | 2 |
| Improved patient access to quality healthcare in remote areas | 17 |
| Improved patient access to healthcare (especially follow-up) | 18 |
| Reduced waiting times for the patient | 3 |
| Physicians' habit of providing in-person consultations does not affect the adoption of telemedicine (they may keep utilising telemedicine even after the ongoing pandemic) | 1 |
| Patients' habit of seeking in-person consultations does not affect the adoption of telemedicine (may utilise telemedicine after the ongoing pandemic) | 1 |
| Less caregiver burden | 6 |
| Reduced stigma in certain clinical specialties | 2 |
| Increased reach for the hospital | 1 |

#### Summary of the barriers to telemedicine systems in India

| **Barriers** | **Count** |
| --- | --- |
| Time-consuming | 3 |
| Financial burden | 12 |
| Need for physical examination and closer observation | 10 |
| Unavailability of technology resources | 14 |
| Inadequate internet connectivity | 11 |
| Poor health/technology literacy (in remote and rural areas) | 16 |
| Patient socioeconomic factors | 1 |
| Demotivated patients | 1 |
| Cultural and language barriers | 11 |
| Lack of awareness and expertise among health professionals | 10 |
| Lack of dedicated and trained professionals | 10 |
| Lack of availability and expertise in telemedicine interventions | 3 |
| Lack of external reward (government initiatives and schemes) | 2 |
| Patients' doubts about the quality of virtual assessment and acceptance | 19 |
| Healthcare providers' beliefs about the efficacy of telemedicine: e.g., not an adequate substitute for in-person evaluations after the pandemic | 5 |
| Resistance to change | 4 |
| Doubts about the ease of using e-consultations | 1 |
| Less family support for telemedicine interventions | 1 |
| Lack of solutions to integrate telemedicine into the existing healthcare system despite overcrowded outpatient services | 5 |
| Patient 'no shows' | 1 |
| Need for social acceptance of telemedicine | 4 |
| Fear of violation of data privacy and security | 20 |
| Technology issues (system installation and maintenance issues) | 8 |
| Practitioner credibility and licensing issues | 1 |
| Legal liability for the healthcare providers | 4 |
| Uncertainty concerning the telemedicine guidelines and their legal ramifications | 16 |

**Summary of selected articles**

| **Author(s) & Year** | **Health system tier** | **Clinical Specialty** | **Type of telemedicine system** | **Population attributes** | **Study perspective** | **Telemedicine Facilitators** | **Telemedicine Barriers** |
| --- | --- | --- | --- | --- | --- | --- | --- |
| Appireddy et al. (2020) | Secondary | Teleneurology | Video Teleconferencing | - | Patients, Doctors | **Patients -**  1. Less caregiver burden  2. Less commute  3. Economical  4. Remote healthcare /convenient service  5. Time-saving  6. Lesser chances of contracting hospital-acquired infections  7. Convenient for differently-abled  **Doctors -**  1. Ease of consultation  2. Reduced budgets | **Patients -**  1. Technology issues  2. Unavailability of technology resources  3. Patients' beliefs about the efficacy of telemedicine  **Doctors -**  1. Need for physical health assessment and closer observation |
| Ateriya et al. (2018) | Primary | Telemedicine | - | - | Society | 1. Remote healthcare /convenient service  2. Improved patient access to healthcare  3. Convenient for differently-abled  4. Economical/Reduced budgets  5. Time-saving  6. Improved patient-provider relationship | 1. Need for physical health assessment and closer observation  2. Uncertainty concerning the Telemedicine guidelines and their legal ramifications  3. Financial burden for the patients  4. Data security and privacy issues |
| Bairapareddy et al. (2021) | Secondary | Telerehabilitation | Mobile Teleconsultation | **Healthcare providers (n=52) -** Physiotherapists, respiratory care therapists, pulmonary care physicians and rehab nurses  Young adults and middle-aged (29 - 41 years) with 5 to 7 years of experience  **Patients (n=30) -** middle-aged (41 - 67 years), undergraduates and knew about telerehabilitation for a year | Healthcare providers, Patients | **Patients -**  1. Time-saving  2. Ease of consultation  3. Less commute  4. Less caregiver burden for rehab sessions  5. Minimal equipment  6. Safe environment | **Healthcare providers -**  1. Poor health literacy  2. Financial burden  3. Demotivated patients  4. Lack of technology/unavailability of resources  5. Lack of awareness among health professionals  6. Lack of external reward (government initiatives and schemes)  7. Lack of dedicated and trained professionals  8. Patients' beliefs about the efficacy of telemedicine  9. Time-consuming  **Patients -**  1. Time-consuming  2. Lack of availability and expertise in telemedicine interventions  3. Language barriers  4. Less family support for telemedicine interventions |
| Bakshi and Tandon (2021b) | Primary | Telemedicine | - | Private and government hospital doctors (n=215)  Mostly government doctors (n=180)  Predominantly young adults and middle-aged (18 - 45 years)  Qualified with MBBS, MD, BAMS | Doctors | 1. Financial risk did not affect the adoption of telemedicine.  2. Improved patient access to healthcare  3. Access to high-speed internet and other technology at the healthcare facilities | 1. Social risk  2. Time-consuming (Time risk)  3. Security and privacy risk  4. Technology risk (Installation and maintenance)  5. Resistance to change |
| Bali (2019) | Primary | Telemedicine | - | - | Society | 1. Easy access/Improved patient access to healthcare  2. Remote healthcare /convenient service  3. Economical/Reduced budgets  4. Less commute  5. Reduced hospital visits for patients  6. Time-saving  7. Safe and effective | 1. Uncertainty concerning the Telemedicine guidelines and their legal ramifications.  2. Data security and privacy issues  3. Lack of guidelines to integrate telemedicine into the existing healthcare system  4. Lack of availability and expertise in TM interventions  5. Lack of dedicated and trained professionals  6. Lack of awareness among health professionals  7. Expensive technology  8. Technology issues  9. Financial burden for the health professionals  10. Unavailability of ICT resources  11. Poor internet connectivity  12. Practitioner credibility and licensing issues  13. Cultural and Language barriers  14. Doubts about the quality of virtual assessment  15. Need for physical health assessment and closer observation  16. Patients' beliefs about the efficacy of telemedicine  17. Poor technology literacy of the patients  18. Resistance to change  19. Legal liability for the healthcare providers |
| Banerjee et al. (2020) | Primary | Dementia Care | Teleconsultation | Healthcare providers (n=148) - Psychiatrists, Neurologists, and General physicians (39.2 ± 5.3 years with mean experience of 10.2 ± 2.4 years) | Doctors | 1. Patients can easily consult online, requiring lesser hospital visits.  2. Improved patient access to healthcare.  3. Simultaneous and easy delivery of information and instructions to several caregivers. | 1. Doubts about the quality of virtual assessment  2. Uncertainty concerning the Telemedicine guidelines and their legal ramifications.  3. Low e-health knowledge among the patients.  4. Need for physical health assessment. |
| Biswas and Batra (2020) | Secondary | Teleophthalmology | - | - | Society | 1. Remote healthcare/convenient service  2. Reduced hospital visits for patients | 1. Uncertainty concerning the Telemedicine guidelines and their legal ramifications  2. poor internet connectivity  3. Poor technology literacy  4. Financial burden for the health professionals  5. Data security and privacy issues  6. Doubts about the efficacy of virtual assessment |
| Chandwani and Kumar (2018) | Tertiary | Surgical Endocrinology | Video Teleconferencing | **Patients & Caregivers -**  thyroid cancer patients from Orissa  22% population in Orissa - poor, low-literate  **Healthcare Providers -**  5 super-specialists and resident doctors at the apex hospital  8 TM coordinators and support staff at the apex hospital  3 surgeons and physicians at the local medical college  4 coordinators and support staff at the local medical college  Experience - 3 to 7 years | Patients & Caregivers, Healthcare Providers | **Healthcare Providers -**  1. Improved patient access to healthcare  2. Remote healthcare  3. Economical  4. Time-saving  5. Less commute  6. acceptable and effective  7. Reduced hospital visits for patients  **Patients & Caregivers -**  1. acceptable and convenient | 1. Cultural and Language barriers  2. unavailability of resources/poor internet connectivity/Technology issues in rural areas  3. Lack of dedicated and trained professionals  4. Patients' beliefs about the efficacy of telemedicine  5. Financial burden for the healthcare providers  **Patients & Caregivers -**  1. Cultural and Language barriers  2. Need for social acceptance of telemedicine  **Healthcare Providers -**  1. Integrating telemedicine into the existing healthcare system despite overcrowded OP services  2. Patient 'no shows' |
| Corley (2018) | Secondary | Telemedicine for stroke care | Mobile Teleconsultation |  | Doctors | 1. Safe and effective  2. Remote healthcare | 1. Data security and privacy issues  2. Lack of expertise among existing medical providers.  3. Cultural and language barriers.  4. Patients' beliefs about the efficacy of telemedicine  5. Not an adequate substitute for in-person evaluations |
| Das et al. (2020) | Primary | Telemedicine | - | Doctors (n=208)  mostly resident doctors (n=162)  The majority were medical graduates (n=111), and almost half were postgraduates (n=97) Substantial percentage of psychiatrists (22%) | Doctors | 1. Reduced waiting times for the patient.  2. Reduced budgets.  3. Patient access to quality healthcare in remote areas. | 1. Data security and privacy issues  2. Need for physical examination and closer observation  3. Sceptical about adopting telemedicine over in-person care in the near future. |
| Dash and Sahoo (2021b) | Primary | Telemedicine | Teleconsultation | Doctors (n=337)  male (55.49%), female (44.51%)  Age group -  (36.8%) - below 30 years  (34.42%) - 30 to 45 years  (28.78%) - above 45 years  Qualification -  Mostly MBBS, followed by MD and MS  Experience -  (38.28%) - up to 5 years  (33.53%) - 5 to 10 years  (28.19%) - more than 10 years  e-consultation Experience -  (51.63%) - less than 1 year  (34.12%) - 1 and 5 years  (14.24%) - more than 5 years | Doctors | 1. Financial risk did not affect the adoption of telemedicine.  2. Physicians' habit of providing in-person consultations did not affect the adoption of telemedicine, and they may continue to use telemedicine after the pandemic.  3. Enables patients to have virtual 24*7 consultation globally at a lower cost | 1. Doubts about the efficacy of virtual consultations are the main barrier  2. Social risk  3. Doubts about technology/availability of resources  4. Doubts about expertise in administering e-consultations |
| Dash and Sahoo (2021a) | Primary | Telemedicine | Teleconsultation | Patients (n=462)  (55.84%) - Male  (44.16%) - Female  Age group -  (27.27%) - below 25 years  (25.76%) - 26 and 35 years  (26.41%) - 36 and 45 years  (20.56%) - above 45 years  Qualification -  Mostly Graduates (n=178), followed by Postgraduates (n=142), Undergraduates (n=74) and others | Patients | 1. Patients' habit of seeking in-person consultations does not affect the adoption of telemedicine (may continue to use telemedicine after the pandemic)  2. Ease of consultation  3. Lesser chances of contracting hospital-acquired infections  4. Improved patient access to healthcare | 1. Costs associated with the e-consultation system are the main barrier  2. Social risk  3. Doubts about technology/availability of resources for e-consultations  4. Doubts about the ease of using e-consultations |
| Galagali et al. (2021) | Secondary | Telepaediatrics (Child and Adolescent Healthcare) | Video Teleconferencing | - | Caregivers, Patients | 1. Remote healthcare /convenient service  2. acceptable, convenient, and effective  3. Reduced hospital visits | 1. illiteracy  2. Fear of violation of data privacy and security  3. Inadequate internet connectivity  4. Need for physical examination and closer observation  5. Uncertainty concerning the Telemedicine guidelines and their legal ramifications. |
| Ghai (2020) | Secondary | Teledentistry | Teleconsultation | - | Doctors, Patients | **Dentists & Patients -**  1. Reduced hospital visits for patients  2. Lesser waiting time for patients  3. Economical | **Dentists -**  1. Lack of external reward (government initiatives and schemes)  2. Doubts about technology/availability of resources  3. Doubts about expertise in administering TM consultations  4. Costs associated with TM systems  5. Doubts about the quality of virtual assessment  6. Uncertainty concerning the Telemedicine guidelines and their legal ramifications.  **Patients -**  1. Patients' beliefs about the efficacy of telemedicine and acceptance |
| Hameed et al. (2021) | Tertiary | Teleurology | Mobile Teleconsultation (Smartphone application) | 33 patients (mean age -47.8 years) | Patients | 1. Patients can easily consult online, requiring lesser hospital visits.  2. Safe and effective  3. Remote healthcare /convenient service | 1. Lack of technical knowledge among the patients  2. Language barriers  3. Poor internet connectivity |
| Iyengar et al. (2020) | Primary | Telemedicine | - | - | Doctors | 1. Ease of consultation | 1. Lack of dedicated and trained professionals  2. Uncertainty concerning the Telemedicine guidelines and their legal ramifications  3. Patient data security and privacy issues  4. Legal liability for the healthcare providers |
| Jayarajan et al. (2020) | Tertiary | Telerehabilitation | Teleconsultation | - | Society | 1. Time-saving  2. Less commute  3. Improved patient access to healthcare.  4. Safe and effective  5. Reduced budgets | 1. Lack of trained professionals  2. Need for physical health assessment.  3. Data security and privacy issues  4. Ambiguity related to the telemedicine guidelines  5. Lack of technology literacy  6. Lack of guidelines to integrate telemedicine into the existing healthcare system with overcrowded outpatient services. |
| Joshi et al. (2021) | Primary | Telemedicine | - | ASHA (Accredited Social Health Activist) workers, nodal officers, laboratory technicians, pharmacists, inventory managers, data entry operators and ambulance operators | Health workers and staff | 1. Remote healthcare  2. Reduced budgets  3. Improved patient access to healthcare. | 1. Lack of dedicated staff  2. Network and connectivity issues in the rural areas  3. Lack of expertise  4. Low e-health knowledge among the patients.  5. Lack of solutions to integrate telemedicine into the existing healthcare system with overcrowded outpatient services. |
| Kalaivanan et al. (2021) | Primary | Telepsychiatry (Geriatric Psychiatry) | Teleconsultation | - | Society, Patients, Caregivers | 1. Economical  2. Time-saving  3. Less commute  **Patients -**  1. Ease of consultation  2. Easy access  **Caregivers -**  1. Less caregiver burden for rehab sessions | 1. Patients' beliefs about the efficacy of telemedicine  2. Lack of dedicated and trained professionals  3. Financial burden for healthcare professionals  4. Data security and privacy issues  5. Lack of technology /unavailability of resources  6. Uncertainty concerning the Telemedicine guidelines and their legal ramifications.  7. Fear of violation of data privacy and security  8. Need for physical examination and closer observation  9. Lack of technology among the patients. |
| Kanuri et al. (2020) | Primary | Telepsychiatry (Cognitive behavioural therapy (CBT) - Generalised anxiety disorder (GAD)) | e-Health (Digital mental intervention) | Students (n=15)  Male (n=13)  Female (n=2)  18 to 22 years  60% - the first year of college  Everyone had good English proficiency | Patients | 1. Acceptable and effective  2. User-friendly technology  3. Reduced stigma and cost  4. Convenient  5. Feasible | 1. Data security and privacy risk  2. Time-consuming  3. Technology issues |
| Kavadichanda et al. (2021) | Tertiary | Telerheumatology | Mobile Teleconsultation | **Patients (n=373) -**  Age - 25 to 44 years  334 patients - economically poorer patients (median monthly household income is less than INR 14000)  122 patients - Families could not afford necessities  **Healthcare providers (n=12) -** Rheumatology consultants (n=2), Rheumatology fellows (n=6), Medical officers (n=2), Specialty nurses (n=2) | Patients, Healthcare providers | **Patients -**  1. Acceptable and cost-effective  **Healthcare providers -**  1. Reduced budgets  2. Ease of consultation | **Healthcare providers -**  Ambiguity related to the telemedicine guidelines |
| Kumar et al. (2019) | Primary | Tuberculosis Care | Mobile Teleconsultation | Patients (n=185)  Age - 35.25 ± 11.59 years  114 patients - Males  121 patients - resided in an urban area  151 patients - used a mobile phone. Only 41 patients communicate with their doctors through their mobile phones. | Patients | 1. Ease of consultation  2. Economical  3. Safe and effective  4. Remote healthcare /convenient service  5. Time-saving  6. Less commute | 1. Poor internet connectivity  2. Data security and privacy issues  3. Cultural and Language barriers |
| Mahajan et al. (2020) | Secondary | Telepaediatrics | - | **-** | Doctors, Patients | **Doctors -**  1. Reduced budgets  2. Less commute  3. Reduced hospital visits for patients  **Patients -**  1. Economical  2. Less commute  3. Reduced hospital visits | **Doctors -**  1. Need for physical health assessment and closer observation  2. Uncertainty concerning the Telemedicine guidelines and their legal ramifications  3. Legal liability for the healthcare providers  4. Patient data security and privacy issues  5. Technology issues  6. Poor technology literacy of the patients  7. Patient socioeconomic factors |
| Mahmood et al. (2019) | Tertiary | Telerehabilitation (Stroke rehabilitation) | Mobile Teleconsultation | **Patients (n=50) -**  Age - 55.2 ± 13.39 years  Male - 36  Female - 14  **Caregivers (n=52) -**  Age - 39.78 ± 12.62 years  Male - 20  Female - 32 | Patients, Caregivers | 1. Acceptable and effective  2. Economical  3. Improved patient access to healthcare (especially follow-up)  4. Time-saving  5. Quick delivery of health information to doctors | 1. Patients' beliefs about the efficacy of telemedicine  2. Data security and privacy issues |
| Manglani et al. (2020) | Secondary | Telepaediatrics (Paediatric HIV Care) | Video Teleconferencing | 2608 children from 28 non-TM centres  2803 children from 31 TM centres  31 TM centres - 12 irregular centres (few (less than 12) TM consultations), 19 regular centres  1365 children - irregular consultations  1438 children - regular TM consultations  Age - Up to 18 years  32% - 10 to 14.9 years  In TM centres, a higher proportion of males compared with females | Patients | 1. Better patient care (few deaths and regular follow-ups)  2. acceptable and effective  3. Economical  4. Enhanced access to healthcare  5. Reduced waiting times  6. Higher patient satisfaction  7. Less commute | 1. Patients' beliefs about the efficacy of telemedicine  2. Healthcare providers' beliefs about the efficacy of telemedicine  3. Lack of dedicated and trained professionals |
| Mittal and Pareek (2020) | Primary | Triaging | Mobile Teleconsultation | 2477 patients (Teleconsultation)  10,625 patients (Mobile Teleconsultation)  29% OP consultation | Patients, Doctors | 1. Lesser chances of contracting hospital-acquired infections for healthcare providers and patients.  2. Reduced hospital visits for patients. | 1. Poor technology literacy in rural areas  2. unavailability of resources  3. Resistance to change |
| Naik et al. (2020) | Primary | Telepsychiatry | Teleconsultation | - | Society | 1. Acceptable and effective  2. Economical  3. Time-saving  4. Improved patient access to healthcare  5. Less caregiver burden  6. Reduced hospital visits for patients  7. Reduced stigma | 1. Cultural and Language barriers  2. Patients' beliefs about the efficacy of telemedicine and acceptance  3. Data security and privacy issues  4. Legal liability for the healthcare providers  5. Lack of technology literacy |
| Ramakrishnan et al. (2020) | Tertiary | Tele-ICU | - | - | Society | 1. Improved patient access to healthcare  2. Reduced caregiver burden  3. Reduced hospital visits for patients  4. Remote healthcare /convenient service | 1. Patients' beliefs about the efficacy of telemedicine  2. Financial burden for the health professionals  3. Uncertainty concerning the Telemedicine guidelines and their legal ramifications.  4. Poor internet connectivity |
| Sharma et al. (2020) | Secondary | Teleophthalmology | Mobile Teleconsultation (Smartphone application) | Ophthalmologists (n=1180) | Healthcare providers, Patients | **Healthcare providers -**  1. Improved patient access to healthcare  2. Remote healthcare /convenient service  **Patients -**  1. Remote healthcare /convenient service  2. Time-saving  3. Less commute  4. Reduced hospital visits  5. Ease of consultation | **Healthcare providers & Patients -**  1. Lack of awareness  2. Doubts about the quality of virtual assessment  3. poor internet connectivity  4. Patients' & Healthcare providers' beliefs about the efficacy of telemedicine  **Healthcare providers -**  1. Financial burden  2. Lack of dedicated and trained professionals  3. Technology issues  4. Uncertainty concerning the Telemedicine guidelines and their legal ramifications.  5. unavailability of resources  6. Data security and privacy issues  **Patients -**  1. Need for physical health assessment and closer observation  2. Poor technology literacy of the patients  3. Expensive technology  4. Cultural and Language barriers  5. Data security and privacy issues |
| Sharma and Prashar (2019) | Primary | Telemedicine | e-Health | - | Patients, Doctors, Organisation, Government | **Patients -**  1. Economical  2. Time-saving  3. Less caregiver burden  **Doctors -**  1. Access to quality healthcare in remote areas  2. Reduced possibility of data omission  **Organisation -**  1. Reduced treatment costs  2. Enhanced safety for the patients and healthcare providers  3. Improved patient care  4. Improved diagnostic and treatment capabilities  5. Increased reach for the hospital  **Government -**  1. Enhanced access to healthcare  2. Cost savings from healthcare delivery  3. Access to quality healthcare in remote areas | **Patients -**  1. Data security and privacy risk  2. Lack of health/technology literacy  3. Doubts about the quality of virtual assessment  **Doctors -**  1. Lack of dedicated and trained professionals  2. Lack of awareness and expertise among health professionals  **Organisation -**  1. Lack of dedicated and trained professionals  2. Resistance to change  3. Perceived financial burden for the healthcare facilities  **Government -**  1. Lack of technology /unavailability of resources  2. Poor internet connectivity  3. Costs associated with the system  4. Lack of technology literacy in remote and rural areas  5. Lack of health literacy  6. Legal complications from the system  7. Cultural and Language barriers |
| Thomas et al. (2020) | Secondary | Tuberculosis Care | mHealth (non-smartphone-based technology) | **Patients (n=62) -**  62 patients with TB  36 - men  30 - HIV coinfected  Age - 18 to 65 years (median  35 years)  Median monthly household income - 8500 INR  **Healthcare Providers (n=31) -**  13 - men  Age - 25 to 56 years (median 38 years) | Patients, healthcare providers | **Healthcare providers -**  (High acceptability)  1. Better healthcare quality  2. Efficient healthcare delivery  3. Improved patient-provider relationship  **Patients -**  1. Reduced hospital visits | **Patients -**  (Variable and negative acceptability)  1. Lack of technology  2. Patients' beliefs about the efficacy of telemedicine  a. Some patients felt that intervention improved the patient-provider relationship  b. Some patients felt insulated by the reduced face-to-face contact with providers  **Healthcare providers -**  1. Increased workload |
| Vadlamani et al. (2020) | Primary | Telepsychiatry | Video Teleconferencing | - | Patients, Healthcare Providers | **Patients -**  1. Acceptable and effective  2. Remote healthcare /convenient service  3. Improved patient access to healthcare  4. Time-saving  5. Cost-effective (underserved population, restricted access population)  **Healthcare Providers -**  Economic/Reduced budgets | **Patients -**  1. Poor internet connectivity  2. Costs associated with the system  3. Patients' beliefs about the efficacy of telemedicine  4. Patient data privacy and security issues  5. Age and disability-related challenges  6. Lack of health/technology literacy  **Healthcare Providers -**  1. Financial burden for the health professionals  2. Uncertainty concerning the Telemedicine guidelines and their legal ramifications  3. Doubts about the quality of virtual assessment  4. Lack of awareness and expertise among health professionals |
| Verma et al. (2022) | Tertiary | Telehepatology | Mobile Teleconsultation | Patients (n=210)  Median age - 46 (35-56) years  32.3% females  70% - belonged to the middle or lower socioeconomic class  61% - from rural areas  109 patients - follow-ups  101 patients - consulted for the first time  Healthcare workers were the common sources of referral (51.4%), followed by patient relatives (24.3%) for teleconsultations | Patients | 1. Economical/Reduced budgets  2. Time-saving  3. Less commute  4. Reduced hospital visits  5. Cost did not affect the adoption of telemedicine.  6. Technology issues and language barriers did not affect the adoption of telemedicine.  7. Improvement in patient's mental and physical quality of life. | 1. Not an adequate substitute for in-person evaluations after the pandemic  2. Poor internet connectivity  3. Data security and privacy issues  4. Cultural and Language barriers |
| Verma et al. (2021) | Primary | Telemedicine | - | - | Society | 1. Reduced budgets  2. Remote healthcare /convenient service  3. Improved patient access to healthcare  4. Time-saving  5. Less commute  6. Reduced hospital visits for patients | 1. Unavailability of resources  2. Poor technology literacy  3. Uncertainty concerning the Telemedicine guidelines and their legal ramifications  4. Data security and privacy issues  5. Cultural and Language barriers  6. Doubts about the quality of virtual assessment  7. Lack of dedicated and trained professionals  8. Technology issues |

#### Facilitators categorised for the three healthcare tiers

| **Factors** | **Facilitators** | **No. of articles** | | |
| --- | --- | --- | --- | --- |
|  |  | **Primary** | **Secondary** | **Tertiary** |
| **Infrastructural** | **Time-saving** | 8 | 3 | 4 |
|  | **Less commute** | 4 | 5 | 3 |
|  | **Reduced hospital visits for patients Reduced hospital visits for patients** | 5 | 6 | 4 |
|  | **Ease of consultation** | 7 | 3 | 3 |
|  | Access to high-speed internet and other technology at the healthcare facilities | 1 |  |  |
|  | Requires only minimal equipment/User-friendly technology | 1 | 1 |  |
|  | Reduced waiting times for the patient | 1 | 2 |  |
| **Socio-Cultural** | Lesser chances of contracting hospital-acquired infections | 2 | 1 |  |
|  | Convenient for differently-abled | 1 | 1 |  |
|  | The financial risk did not affect the adoption of telemedicine | 2 |  | 1 |
|  | Technology issues and language barriers did not affect the adoption of telemedicine |  |  | 1 |
|  | **Safe, acceptable, and effective** | **6** | **4** | **5** |
|  | Higher patient satisfaction |  | 1 |  |
|  | Improved patient's mental and physical quality of life |  |  | 1 |
|  | Physicians' habit of providing in-person consultations does not affect the adoption of telemedicine (they may continue to use telemedicine after the pandemic) | 1 |  |  |
|  | Patients' habit of seeking in-person consultations does not affect the adoption of telemedicine (they may continue to use telemedicine after the pandemic) | 1 |  |  |
|  | **Less caregiver burden** | 3 | 2 | 1 |
|  | Reduced stigma in certain clinical specialties | 2 |  |  |
| **Regulatory & Financial** | **Reduced healthcare costs Reduced healthcare costs** | **11** | **4** | **5** |
|  | Reduced possibility of medical data omission | 1 |  | 1 |
|  | Improved efficiency and quality of care | 1 | 2 |  |
|  | Improved patient-provider relationship | 1 | 1 |  |
|  | **Improved patient access to quality healthcare in remote areas** | 8 | 5 | 3 |
|  | **Improved patient access to healthcare Improved patient access to healthcare** | 12 | 2 | 4 |
|  | Simultaneous and easy delivery of information and instructions to several caregivers | 1 |  |  |
|  | Enhanced safety for the patients and healthcare providers | 1 |  |  |
|  | Increased reach for the hospital | 1 |  |  |

#### Barriers categorised for the three healthcare tiers

| **Factors** | **Barriers** | **No. of articles** | | |
| --- | --- | --- | --- | --- |
|  |  | **Primary** | **Secondary** | **Tertiary** |
| **Infrastructural** | Time-consuming | 2 | 2 |  |
|  | **Unavailability of technology resources** | 7 | 5 |  |
|  | **Inadequate internet connectivity** | 4 | 3 | 4 |
|  | Technology issues (system installation and maintenance issues) | 4 | 3 |  |
|  | Lack of awareness and expertise among health professionals | 3 | 4 |  |
|  | **Lack of dedicated and trained professionals** | 6 | 3 | 2 |
|  | Lack of availability and expertise in telemedicine interventions | 3 | 1 |  |
| **Socio-Cultural** | **Need for physical examination and closer observation** | **5** | **4** | **1** |
|  | **Poor health/technology literacy (in remote and rural areas)** | 9 | 5 | 2 |
|  | Patient socioeconomic factors |  | 1 |  |
|  | Demotivated patients |  | 1 |  |
|  | **Cultural and language barriers** | 5 | 3 | 3 |
|  | Need for social acceptance of telemedicine | 3 |  | 1 |
|  | **Patients' doubts about the quality of virtual assessment and acceptance** | **7** | **8** | **3** |
|  | **Healthcare providers' beliefs about the efficacy of telemedicine - Not an adequate substitute for in-person evaluations after the pandemic** | **2** | **3** | **1** |
|  | Resistance to change | 4 |  |  |
|  | Doubts about the ease of using e-consultations | 1 |  |  |
|  | Less family support for telemedicine interventions |  | 1 |  |
| **Regulatory & Financial** | Lack of solutions to integrate telemedicine into the existing healthcare system (despite overcrowded outpatient services) | 2 |  | 2 |
|  | Patient 'no shows' |  |  | 1 |
|  | **Financial burden** | **6** | **4** | **2** |
|  | **Fear of violation of data privacy and security** | 12 | 5 | 3 |
|  | Practitioner credibility and licensing issues | 1 |  |  |
|  | Legal liability for the healthcare providers | 3 | 1 |  |
|  | **Uncertainty concerning the telemedicine guidelines and their legal ramifications** | 8 | 5 | 3 |
|  | Lack of external reward (government initiatives and schemes) |  | 2 |  |

**Barriers and facilitators categorised by the delivery modes**

| **Mode of Telemedicine Delivery** | | **Facilitators** | **Barriers** |
| --- | --- | --- | --- |
| **Tele/Video** | **Teleconsultation** | 1. Ease of consultation 2. Economical 3. Improved patient access to healthcare 4. Reduced hospital visits for patients | 1. Unavailability of technology resources 2. Need for social acceptance of telemedicine 3. Patients’ doubts about the quality of virtual assessment and acceptance 4. Legal liability for healthcare providers |
|  | **Video Teleconferencing** | 1. Economical 2. Time-saving 3. Less commute 4. Acceptable and effective 5. Improved patient access to healthcare 6. Remote healthcare/convenient service | 1. Patients’ doubts about the quality of virtual assessment and acceptance 2. Lack of dedicated and trained professionals 3. Unavailability of technology resources 4. Fear of violation of data privacy and security |
| **mHealth** | **Mobile Teleconsultation** | 1. Economical 2. Time-saving 3. Less commute 4. Remote healthcare/convenient service 5. Safe, acceptable, and effective 6. Ease of consultation 7. Reduced hospital visits for patients | 1. Inadequate internet connectivity 2. Unavailability of technology resources 3. Poor health/technology literacy 4. Fear of violation of data privacy and security 5. Cultural and language barriers 6. Doubts about the quality of virtual assessment and acceptance 7. Lack of dedicated and trained professionals 8. Uncertainty concerning the telemedicine guidelines and their legal ramifications |
|  | **Non-smartphone-based technology** | 1. Enhanced quality of care and effectiveness 2. Improved patient-provider relationship 3. Reduced hospital visits for patients | 1. Patients’ doubts about the quality of virtual assessment and acceptance 2. Unavailability of technology resources |
