## Supplementary File2 for "Facilitators and Barriers for Telemedicine Systems in India from Multiple Stakeholder Perspectives and Settings"

| S.No. | Citation | Reasons for exclusion |
| --- | --- | --- |
| 1 | Adarmouch, L., Tourari, S., Sebbani, M., & Amine, M. (2021). Impact of the COVID-19 pandemic on the activity of private medical practices in Morocco. Int J Clin Pract, 75(6), e14127. https://doi.org/10.1111/ijcp.14127 | Not focussed on India<br>Outcomes do not address the barriers or facilitators for the adoption and use of telemedicine systems |
| 2 | Adhikari, S. D., Biswas, S., Mishra, S., Kumar, V., Bharti, S. J., Gupta, N., Garg, R., & Bhatnagar, S. (2021). Telemedicine as an Acceptable Model of Care in Advanced stage Cancer Patients in the Era of Coronavirus Disease 2019 - An Observational Study in a Tertiary Care Centre. Indian J Palliat Care, 27(2), 306-312. https://doi.org/10.25259/IJPC_45_21 | 4 (no reasons behind the barriers and facilitators) |
| 3 | Agarwal, N., & Biswas, B. (2020). Doctor Consultation through Mobile Applications in India: An Overview, Challenges and the Way Forward. Healthc Inform Res, 26(2), 153-158. https://doi.org/10.4258/hir.2020.26.2.153 | 4 |
| 4 | Ajay, V. S., Jindal, D., Roy, A., Venugopal, V., Sharma, R., Pawar, A., Kinra, S., Tandon, N., & Prabhakaran, D. (2016). Development of a Smartphone-Enabled Hypertension and Diabetes Mellitus Management Package to Facilitate Evidence-Based Care Delivery in Primary Healthcare Facilities in India: The mPower Heart Project. J Am Heart Assoc, 5(12). https://doi.org/10.1161/JAHA.116.00434[illegible] | Outcomes do not address the barriers or facilitators for the adoption and use of telemedicine systems |
| 5 | Akhtar, N., Rajan, S., Chakrabarti, D., Kumar, V., Gupta, S., Misra, S., Chaturvedi, A., Azhar, T., Parveen, S., Qayoom, S., Niranjan, P., & Tiwari, S. (2021). Continuing cancer surgery through the first six months of the COVID-19 pandemic at an academic university hospital in India: A lower-middle-income country experience [Article]. J Surg Oncol, 123(5), 1177-1187. https://doi.org/10.1002/jso.26419 | Outcomes do not address the barriers or facilitators for the adoption and use of telemedicine systems |
| 6 | Alam, M. N., Munjal, S., Panda, N., Garg, M., & Shipra, M. K. (2021). Efficacy of the Smartphone App for sending Text Reminders to reduce 'No Shows' in Speech Therapy Sessions at a Tertiary Care Centre in India [Article]. Disability, CBR & Inclusive Development, 32(2), 127-141. https://doi.org/10.47985/dcidj.470 | Outcomes do not address the barriers or facilitators for the adoption and use of telemedicine systems |
| 7 | Alrahbi, D. A., Khan, M., Gupta, S., Modgil, S., & Chiappetta Jabbour, C. J. (2020). Challenges for developing health-care knowledge in the digital age. JOURNAL OF KNOWLEDGE MANAGEMENT, ahead-of-print(ahead-of-print). https://doi.org/10.1108/jkm-03-2020-0224 | Not associated with patient health<br>Not focussed on India |
| 8 | Andrees, V., Klein, T. M., Augustin, M., & Otten, M. (2020). Live interactive teledermatology compared to in-person care - a systematic review. J Eur Acad Dermatol Venereol, 34(4), 733-745. https://doi.org/10.1111/jdv.16070 | Not focussed on India |
| 9 | Archer, N., Lokker, C., Ghasemaghaei, M., & DiLiberto, D. (2021). eHealth Implementation Issues in Low-Resource Countries: Model, Survey, and Analysis of User Experience. J Med Internet Res, 23(6), e23715. https://doi.org/10.2196/23715 | Not focussed on India |

| | | |
|---|---|---|
| 10 | Assayag, E., Tsessler, M., Wasser, L. M., Drabkin, E., Reich, E., Weill, Y., Zadok, D., Nair, A. G., & Andron, A. (2021). Telemedicine comes of age during coronavirus disease 2019 (COVID-19): An international survey of oculoplastic surgeons [Article]. Eur J Ophthalmol, 31(6), 2881-2885. https://doi.org/10.1177/1120672120965471 | Not focussed on India<br>Outcomes do not address the barriers or facilitators for the adoption and use of telemedicine systems |
| 11 | Barigela, R., Kodali, P. B., & Hense, S. (2021). What is Stopping Primary Health Centers to Go Digital? Findings of a Mixed-method Study at a District Level Health System in Southern India. Indian J Community Med, 46(1), 97-101. https://doi.org/10.4103/ijcm.IJCM_304_20 | Not associated with patient health |
| 12 | Bassi, A., John, O., Praveen, D., Maulik, P. K., Panda, R., & Jha, V. (2018). Current Status and Future Directions of mHealth Interventions for Health System Strengthening in India: Systematic Review [Review]. JMIR Mhealth Uhealth, 6(10), e11440, Article e11440. https://doi.org/10.2196/11440 | Outcomes do not address the barriers or facilitators for the adoption and use of telemedicine systems |
| 13 | Bhargava, S., McKeever, C., & Kroumpouzos, G. (2021). Impact of COVID-19 pandemic on dermatology practices: Results of a web-based, global survey [Article]. Int J Womens Dermatol, 7(2), 217-223. https://doi.org/10.1016/j.ijwd.2020.09.010 | Not focussed on India |
| 14 | Bhargava, S., Negbenebor, N., Sadoughifar, R., Ahmad, S., & Kroumpouzos, G. (2021). Global impact on dermatology practice due to the COVID-19 pandemic [Article]. Clin Dermatol, 39(3), 479-487. https://doi.org/10.1016/j.clindermatol.2021.01.017 | Not focussed on India<br>Outcomes do not address the barriers or facilitators for the adoption and use of telemedicine systems |
| 15 | Bhaskar, S., Bradley, S., Chattu, V. K., Adisesh, A., Nurtazina, A., Kyrykbayeva, S., Sakhamuri, S., Yaya, S., Sunil, T., Thomas, P., Mucci, V., Moguilner, S., Israel-Korn, S., Alacapa, J., Mishra, A., Pandya, S., Schroeder, S., Atreja, A., Banach, M., & Ray, D. (2020). Telemedicine Across the Globe-Position Paper From the COVID-19 Pandemic Health System Resilience PROGRAM (REPROGRAM) International Consortium (Part 1) [Article]. Front Public Health, 8, 556720, Article 556720. https://doi.org/10.3389/fpubh.2020.556720 | Not focussed on India |
| 16 | Bhat, A., Goud, B. R., Pradeep, J. R., Jayaram, G., Radhakrishnan, R., & Srinivasan, K. (2020). Can Mobile Health Improve Depression Treatment Access and Adherence Among Rural Indian Women? A Qualitative Study. Cult Med Psychiatry, 44(4), 461-478. https://doi.org/10.1007/s11013-019-09664-3 | 4 (barriers and facilitators are dicussed for adherence to this specific intervention) |
| 17 | Bhatt, S., Evans, J., & Gupta, S. (2018). Barriers to Scale of Digital Health Systems for Cancer Care and Control in Last-Mile Settings. J Glob Oncol, 4, 1-3. https://doi.org/10.1200/JGO.2016.007179 | Not focussed on India |
| 18 | Bhatt, S., Isaac, R., Finkel, M., Evans, J., Grant, L., Paul, B., & Weller, D. (2018). Mobile technology and cancer screening: Lessons from rural India. J Glob Health, 8(2), 020421. https://doi.org/10.7189/jogh.08.020421 | Outcomes do not address the barriers or facilitators for the adoption and use of telemedicine systems |

| | | |
|---|---|---|
| 19 | Birur, N. P., Patrick, S., Bajaj, S., Raghavan, S., Suresh, A., Sunny, S. P., Chigurupati, R., Wilder-Smith, P., Gurushanth, K., Gurudath, S., Rao, P., & Kuriakose, M. A. (2018). A Novel Mobile Health Approach to Early Diagnosis of Oral Cancer. J Contemp Dent Pract, 19(9), 1122-1128. https://www.ncbi.nlm.nih.gov/pubmed/30287715 | Not concerned with patient monitoring, screening, diagnostic tests, or treatment |
| 20 | Biswas, S., Adhikari, S. D., & Bhatnagar, S. (2020). Integration of Telemedicine for Home-Based End-of-Life Care in Advanced Cancer Patients during Nationwide Lockdown: A Case Series. Indian J Palliat Care, 26(Suppl 1), S176-S178. https://doi.org/10.4103/IJPC.IJPC_174_20 | Outcomes do not address the barriers or facilitators for the adoption and use of telemedicine systems |
| 21 | Bucher, S. L., Cardellichio, P., Muinga, N., Patterson, J. K., Thukral, A., Deorari, A. K., Data, S., Umoren, R., & Purkayastha, S. (2020). Digital Health Innovations, Tools, and Resources to Support Helping Babies Survive Programs. Pediatrics, 146(Suppl 2), S165-S182. https://doi.org/10.1542/peds.2020-016915I | Not focussed on India |
| 22 | Budd, J., Miller, B. S., Manning, E. M., Lampos, V., Zhuang, M., Edelstein, M., Rees, G., Emery, V. C., Stevens, M. M., Keegan, N., Short, M. J., Pillay, D., Manley, E., Cox, I. J., Heymann, D., Johnson, A. M., & McKendry, R. A. (2020). Digital technologies in the public-health response to COVID-19. Nat Med, 26(8), 1183-1192. https://doi.org/10.1038/s41591-020-1011-4 | Not associated with patient health<br>Not focussed on India |
| 23 | Burnand, H. G., McMahon, S. E., Sayers, A., Tshengu, T., Gibson, N., Blom, A. W., Whitehouse, M. R., & Wylde, V. (2021). The EQ-5D-3L administered by text message compared to the paper version for hard-to-reach populations in a rural South African trauma setting: a measurement equivalence study. Arch Orthop Trauma Surg, 141(6), 947-957. https://doi.org/10.1007/s00402-020-03574-5 | Not focussed on India |
| 24 | Burton, M. J., Ramke, J., Marques, A. P., Bourne, R. R. A., Congdon, N., Jones, I., Ah Tong, B. A. M., Arunga, S., Bachani, D., Bascaran, C., Bastawrous, A., Blanchet, K., Braithwaite, T., Buchan, J. C., Cairns, J., Cama, A., Chagunda, M., Chuluunkhuu, C., Cooper, A., . . . Faal, H. B. (2021). The Lancet Global Health Commission on Global Eye Health: vision beyond 2020 [Review]. Lancet Glob Health, 9(4), e489-e551. https://doi.org/10.1016/S2214-109X(20)30488-5 | Not focussed on India<br>Outcomes do not address the barriers or facilitators for the adoption and use of telemedicine systems |
| 25 | Carmichael, S. L., Mehta, K., Srikantiah, S., Mahapatra, T., Chaudhuri, I., Balakrishnan, R., Chaturvedi, S., Raheel, H., Borkum, E., Trehan, S., Weng, Y., Kaimal, R., Sivasankaran, A., Sridharan, S., Rotz, D., Tarigopula, U. K., Bhattacharya, D., Atmavilas, Y., Pepper, K. T., . . . Ananya Study, G. (2019). Use of mobile technology by frontline health workers to promote reproductive, maternal, newborn and child health and nutrition: a cluster randomized controlled Trial in Bihar, India. J Glob Health, 9(2), 0204249. https://doi.org/10.7189/jogh.09.020424 | Not concerned with patient monitoring, screening, diagnostic tests, or treatment |

| | | |
|---|---|---|
| 26 | Chan, R. J., Crichton, M., Crawford-Williams, F., Agbejule, O. A., Yu, K., Hart, N. H., de Abreu Alves, F., Ashbury, F. D., Eng, L., Fitch, M., Jain, H., Jefford, M., Klemanski, D., Koczwara, B., Loh, K., Prasad, M., Rugo, H., Soto-Perez-de-Celis, E., van den Hurk, C., . . . Multinational Association of Supportive Care in Cancer Survivorship Study, G. (2021). The efficacy, challenges, and facilitators of telemedicine in post-treatment cancer survivorship care: an overview of systematic reviews. Ann Oncol, 32(12), 1552-1570. https://doi.org/10.1016/j.annonc.2021.09.001 | Not focussed on India |
| 27 | Chandrasekaran, S., Chandrashekar, V. S., Dalvie, S., & Sinha, A. (2021). The case for the use of telehealth for abortion in India. Sex Reprod Health Matters, 29(2), 1920566. https://doi.org/10.1080/26410397.2021.1920566 | 4 (This is a proposed intervention) |
| 28 | Chauhan, G., & Thakur, J. (2021). Integrated digital approach for prevention and control of noncommunicable diseases: Review of the progress of “mukhyamantri nirog yojna” in the state of Himachal Pradesh, India. INTERNATIONAL JOURNAL OF NONCOMMUNICABLE DISEASES, 6(1), 4-9. https://doi.org/10.4103/jncd.jncd_70_20 | Outcomes do not address the barriers or facilitators for the adoption and use of telemedicine systems |
| 29 | Chawla, G., Abrol, N., & Kakkar, K. (2020). Personal Protective Equipment: A Pandora's Box. Indian J Crit Care Med, 24(5), 371-372. https://doi.org/10.5005/jp-journals-10071-23443 | Not associated with patient health |
| 30 | Chhabra, H. S., Bagaraia, V., Keny, S., Kalidindi, K. K. V., Mallepally, A., Dhillon, M. S., Malhotra, R., & Rajasekharan, S. (2020). COVID-19: Current Knowledge and Best Practices for Orthopaedic Surgeons [Review]. Indian J Orthop, 54(4), 1-15. https://doi.org/10.1007/s43465-020-00135-1 | Outcomes do not address the barriers or facilitators for the adoption and use of telemedicine systems |
| 31 | Cicinelli, M. V., Marmamula, S., & Khanna, R. C. (2020). Comprehensive eye care - issues, challenges, and way forward. Indian J Ophthalmol, 68(2), 316-323. [illegible] | Outcomes do not address the barriers or facilitators for the adoption and use of telemedicine systems |
| 32 | Coorey, G., Peiris, D., Neubeck, L., & Redfern, J. (2020). A realist evaluation approach to explaining the role of context in the impact of a complex eHealth intervention for improving prevention of cardiovascular disease. BMC Health Serv Res, 20(1), 764. https://doi.org/10.1186/s12913-020-[illegible] | Not focussed on India |
| 33 | Crowder, R., Kityamuwesi, A., Kiwanuka, N., Lamunu, M., Namale, C., Tinka, L. K., Nakate, A. S., Ggita, J., Turimumahoro, P., Babirye, D., Oyuku, D., Berger, C. A., Tucker, A., Patel, D., Sammann, A., Dowdy, D., Stavia, T., Cattamanchi, A., & Katamba, A. (2020). Study protocol and implementation details for a pragmatic, stepped-wedge cluster randomised trial of a digital adherence technology to facilitate tuberculosis treatment completion. BMJ Open, 10(11), e039895. https://doi.org/10.1136/bmjopen-2020-039895 | Not focussed on India |
| 34 | Dash, S., Aarthy, R., & Mohan, V. (2021). Telemedicine during COVID-19 in India-a new policy and its challenges. J Public Health Policy, 42(3), 501-509. https://doi.org/10.1057/s41271-021-00287-w | 4 (no reasons behind the barriers and facilitators) |

| | | |
|---|---|---|
| 35 | Davalbhakta, S., Sharma, S., Gupta, S., Agarwal, V., Pandey, G., Misra, D. P., Naik, B. N., Goel, A., Gupta, L., & Agarwal, V. (2020). Private Health Sector in India-Ready and Willing, Yet Underutilized in the Covid-19 Pandemic: A Cross-Sectional Study [Article]. Front Public Health, 8, 571419, Article 571419. https://doi.org/10.3389/fpubh.2020.571419 | Outcomes do not address the barriers or facilitators for the adoption and use of telemedicine systems |
| 36 | Devi, R., Kanitkar, K., Narendhar, R., Sehmi, K., & Subramaniam, K. (2020). A Narrative Review of the Patient Journey Through the Lens of Non-communicable Diseases in Low- and Middle-Income Countries [Review]. Adv Ther, 37(12), 4808-4830. https://doi.org/10.1007/s12325-020-01519-3 | Not focussed on India |
| 37 | Deshpande, S., Patil, D., Dhokar, A., Bhanushali, P., & Katge, F. (2021). Teledentistry: A Boon Amidst COVID-19 Lockdown-A Narrative Review [Review]. Int J Telemed Appl, 2021, 8859746, Article 8859746. https://doi.org/10.1155/2021/8859746 | 4 (barriers and facilitators are dicussed in the event of a pandemic) |
| 38 | Dhadge, N., Shevade, M., Kale, N., Narke, G., Pathak, D., Barne, M., Madas, S., & Salvi, S. (2020). Monitoring of inhaler use at home with a smartphone video application in a pilot study. NPJ Prim Care Respir Med, 30(1), 46. https://doi.org/10.1038/s41533-020-00203-x | Outcomes do not address the barriers or facilitators for the adoption and use of telemedicine systems |
| 39 | Dinakaran, D., Basavarajappa, C., Manjunatha, N., Kumar, C. N., & Math, S. B. (2020). Telemedicine Practice Guidelines and Telepsychiatry Operational Guidelines, India-A Commentary. Indian J Psychol Med, 42(5 Suppl), 1S-3S. https://doi.org/10.1177/0253717620958382 | 4 (discusses the guidelines for TM and Telepsychiatry) |
| 40 | Dolk, H., Leke, A. Z., Whitfield, P., Moore, R., Karnell, K., Barisic, I., Barlow-Mosha, L., Botto, L. D., Garne, E., Guatibonza, P., Godfred-Cato, S., Halleux, C. M., Holmes, L. B., Moore, C. A., Orioli, I., Raina, N., & Valencia, D. (2021). Global birth defects app: An innovative tool for describing and coding congenital anomalies at birth in low resource settings. Birth Defects Res, 113(14), 1057-1073. https://doi.org/10.1002/bdr2.1898 | Not focussed on India |
| 41 | Dorsey, E. R., Glidden, A. M., Holloway, M. R., Birbeck, G. L., & Schwamm, L. H. (2018). Teleneurology and mobile technologies: the future of neurological care. Nat Rev Neurol, 14(5), 285-297. https://doi.org/10.1038/nrneurol.2018.31 | Not focussed on India |
| 42 | D'Souza, J., Biswas, A., Gada, P., Mangroliya, J., & Natarajan, M. (2021). Barriers leading to increased disability in neurologically challenged populations during COVID-19 pandemic: a scoping review. Disabil Rehabil, 1-14. https://doi.org/10.1080/09638288.2021.1986747 | Not focussed on India |
| 43 | Eichberg, D. G., Basil, G. W., Di, L., Shah, A. H., Luther, E. M., Lu, V. M., Perez-Dickens, M., Komotar, R. J., Levi, A. D., & Ivan, M. E. (2020). Telemedicine in Neurosurgery: Lessons Learned from a Systematic Review of the Literature for the COVID-19 Era and Beyond. Neurosurgery, 88(1), E1-E12. https://doi.org/10.1093/neuros/nyaa306 | Not focussed on India<br>Outcomes do not address the barriers or facilitators for the adoption and use of telemedicine systems |

| | | |
|---|---|---|
| 44 | Faujdar, D. S., Singh, T., Kaur, M., Sahay, S., & Kumar, R. (2021). Stakeholders' Perceptions of the Implementation of a Patient-Centric Digital Health Application for Primary Healthcare in India. Healthc Inform Res, 27(4), 315-324. https://doi.org/10.4258/hir.2021.27.4.315 | 1 (perceptions of stakeholders regarding Electronic Health Record, e-prescriptions, longitudinal tracking of clients, appointment reminders, targeted health education in primary health care information system) |
| 45 | Fleming, K. A., Horton, S., Wilson, M. L., Atun, R., DeStigter, K., Flanigan, J., Sayed, S., Adam, P., Aguilar, B., Andronikou, S., Boehme, C., Cherniak, W., Cheung, A. N., Dahn, B., Donoso-Bach, L., Douglas, T., Garcia, P., Hussain, S., Iyer, H. S., . . . Walia, K. (2021). The Lancet Commission on diagnostics: transforming access to diagnostics [Review]. Lancet, 398(10315), 1997-2050. https://doi.org/10.1016/S0140-6736(21)00673-5 | Not focussed on India |
| 46 | Fottrell, E., Jennings, H., Kuddus, A., Ahmed, N., Morrison, J., Akter, K., Shaha, S. K., Nahar, B., Nahar, T., Haghparast-Bidgoli, H., Khan, A. K., Costello, A., & Azad, K. (2016). The effect of community groups and mobile phone messages on the prevention and control of diabetes in rural Bangladesh: study protocol for a three-arm cluster randomised controlled trial. Trials, 17(1), 600. https://doi.org/10.1186/s13063-016-1738-x | Not focussed on India |
| 47 | Galle, A., Semaan, A., Huysmans, E., Audet, C., Asefa, A., Delvaux, T., Afolabi, B. B., El Ayadi, A. M., & Benova, L. (2021). A double-edged sword-telemedicine for maternal care during COVID-19: findings from a global mixed-methods study of healthcare providers. BMJ Glob Health, 6(2). https://doi.org/10.1136/bmjgh-2020-004575 | Not focussed on India |
| 48 | Garg, D., Majumdar, R., Chauhan, S., Preenja, R., Parihar, J., Saluja, A., & Dhamija, R. K. (2021). Teleneurorehabilitation Among Person with Parkinson's Disease in India: The Initial Experience and Barriers to Implementation [Article]. Ann Indian Acad Neurol, 24(4), 536-541. https://doi.org/10.4103/aian.AIAN_127_21 | 4 |
| 49 | Garg, S., Bhatnagar, N., Singh, M. M., Borle, A., Raina, S. K., Kumar, R., & Galwankar, S. (2020). Strengthening public healthcare systems in India; Learning lessons in COVID-19 pandemic. J Family Med Prim Care, 9(12), 5853-5857. https://doi.org/10.4103/jfmpc.jfmpc_1187_20 | Outcomes do not address the barriers or facilitators for the adoption and use of telemedicine systems |
| 50 | Garg, S., Gangadharan, N., Bhatnagar, N., Singh, M. M., Raina, S. K., & Galwankar, S. (2020). Telemedicine: Embracing virtual care during COVID-19 pandemic. J Family Med Prim Care, 9(9), 4516-4520. https://doi.org/10.4103/jfmpc.jfmpc_918_20 | 3 |
| 51 | Gibbard, M., Ponton, E., Sidhu, B. V., Farrell, S., Bone, J. N., Wu, L. A., Schaeffer, E., Cooper, A., Aroojis, A., Mulpuri, K., & Global Pediatric Orthopaedic, G. (2021). Survey of the Impact of COVID-19 on Pediatric Orthopaedic Surgeons Globally. J Pediatr Orthop, 41(8), e692-e697. https://doi.org/10.1097/BPO.0000000000001887 | Not focussed on India<br>Outcomes do not address the barriers or facilitators for the adoption and use of telemedicine systems |

| | | |
|---|---|---|
| 52 | Gopalakrishnan, L., Buback, L., Fernald, L., Walker, D., Diamond-Smith, N., & in addition to The, C. A. S. E. C. (2020). Using mHealth to improve health care delivery in India: A qualitative examination of the perspectives of community health workers and beneficiaries. PLoS One, 15(1), e0227451. https://doi.org/10.1371/journal.pone.0227451 | 2 (role of intervention in influencing community embeddedness, changing the beneficiary-AWW interactions, and provision of health information) |
| 53 | Green, B. N., Pence, T. V., Kwan, L., & Rokicki-Parashar, J. (2020). Rapid Deployment of Chiropractic Telehealth at 2 Worksite Health Centers in Response to the COVID-19 Pandemic: Observations from the Field. J Manipulative Physiol Ther, 43(5), 404 e401-404 e410. https://doi.org/10.1016/j.jmpt.2020.05.008 | Not focussed on India |
| 54 | Grewal, U. S., Shankar, A., Saini, D., Seth, T., Roy, S., Aden, D., Bhandari, D., & Singh, P. (2021). Tele-health and cancer care in the era of COVID-19: New opportunities in low and middle income countries (LMICs). Cancer Treat Res Commun, 27, 100313. https://doi.org/10.1016/j.ctarc.2021.100313 | Not focussed on India<br>Outcomes do not address the barriers or facilitators for the adoption and use of telemedicine systems |
| 55 | Gudi, N., Konapur, R., John, O., Sarbadhikari, S., & Landry, M. (2021). Telemedicine supported strengthening of primary care in WHO South East Asia region: lessons from the COVID-19 pandemic experiences [Article]. BMJ Innovations, 7(3), 580-585, Article bmjinnov-2021-000699. https://doi.org/10.1136/bmjinnov-2021-000699 | 3 |
| 56 | Gudi, N., Lakiang, T., Pattanshetty, S., Sarbadhikari, S. N., & John, O. (2021). Challenges and prospects in india's digital health journey. Indian J Public Health, 65(2), 209-212. https://doi.org/10.4103/ijph.IJPH_1446_20 | 1 (ImplementatIon challenges of digital knowledge system framework) |
| 57 | Gummidi, B., John, O., & Jha, V. (2020). Continuum of care for non-communicable diseases during COVID-19 pandemic in rural India: A mixed methods study. J Family Med Prim Care, 9(12), 6012-6017. https://doi.org/10.4103/jfmpc.jfmpc_1805_20 | Not focussed on India<br>Outcomes do not address the barriers or facilitators for the adoption and use of telemedicine systems |
| 58 | Gupta, R., Kumar, V. M., Tripathi, M., Datta, K., Narayana, M., Ranjan Sarmah, K., Bhatia, M., Devnani, P., Das, S., Shrivastava, D., Gourineni, R. D., Singh, T. D., Jindal, A., & Mallick, H. N. (2020). Guidelines of the Indian Society for Sleep Research (ISSR) for Practice of Sleep Medicine during COVID-19 [Article]. Sleep Vigil, 4(2), 1-12. https://doi.org/10.1007/s41782-020-00097-2 | 4 (discusses the guidelines for TM in sleep medicine during pandemic) |
| 59 | Himmelfarb, J., Vanholder, R., Mehrotra, R., & Tonelli, M. (2020). The current and future landscape of dialysis. Nat Rev Nephrol, 16(10), 573-585. https://doi.org/10.1038/s41581-020-0315-4 | Not focussed on India<br>Outcomes do not address the barriers or facilitators for the adoption and use of telemedicine systems |
| 60 | Hossain, M. M., Tasnim, S., Sharma, R., Sultana, A., Shaik, A. F., Faizah, F., Kaur, R., Uppuluri, M., Sribhashyam, M., & Bhattacharya, S. (2019). Digital interventions for people living with non-communicable diseases in India: A systematic review of intervention studies and recommendations for future research and development [Review]. Digit Health, 5, 2055207619896153. https://doi.org/10.1177/2055207619896153 | Outcomes do not address the barriers or facilitators for the adoption and use of telemedicine systems |

| | | |
|---|---|---|
| 61 | Hrishi, A. P., Prathapadas, U., Praveen, R., Vimala, S., & Sethuraman, M. (2021). A Comparative Study to Evaluate the Efficacy of Virtual Versus Direct Airway Assessment in the Preoperative Period in Patients Presenting for Neurosurgery: A Quest for Safer Preoperative Practice in Neuroanesthesia in the Backdrop of the COVID-19 Pandemic! J Neurosci Rural Pract, 12(4), 718-725. https://doi.org/10.1055/s-0041-1735824 | Outcomes do not address the barriers or facilitators for the adoption and use of telemedicine systems |
| 62 | Ibrahim, A. E., Magdy, M., Khalaf, E. M., Mostafa, A., & Arafa, A. (2021). Teledermatology in the time of COVID-19. Int J Clin Pract, 75(12), e15000. https://doi.org/10.1111/ijcp.15000 | Not focussed on India |
| 63 | Jawade, P., Hiware, G., Reche, A., Madhu, P. P., Chhabra, K. G., & Kitey, V. (2021). New Understanding of Dental Public Health: A Review. JOURNAL OF PHARMACEUTICAL RESEARCH INTERNATIONAL, 33(50B), 72-78. https://doi.org/10.9734/jpri/2021/v33i50B33429 | Outcomes do not address the barriers or facilitators for the adoption and use of telemedicine systems |
| 64 | Jayadev, C., Mahendradas, P., Vinekar, A., Kemmanu, V., Gupta, R., Pradhan, Z. S., D'Souza, S., Aroor, C. D., Kaweri, L., Shetty, R., Honavar, S. G., & Shetty, B. (2020). Tele-consultations in the wake of COVID-19 - Suggested guidelines for clinical ophthalmology. Indian J Ophthalmol, 68(7), 1316-1327. https://doi.org/10.4103/ijo.IJO_1509_20 | 4 (discusses the guidelines for TM in ophthalmology) |
| 65 | Jeemon, P., Severin, T., Amodeo, C., Balabanova, D., Campbell, N. R. C., Gaita, D., Kario, K., Khan, T., Melifonwu, R., Moran, A., Ogola, E., Ordunez, P., Perel, P., Pineiro, D., Pinto, F. J., Schutte, A. E., Wyss, F. S., Yan, L. L., Poulter, N. R., & Prabhakaran, D. (2021). World Heart Federation Roadmap for Hypertension - A 2021 Update. Glob Heart, 16(1), 63. https://doi.org/10.5334/gh.1066 | Not focussed on India<br>Outcomes do not address the barriers or facilitators for the adoption and use of telemedicine systems |
| 66 | Jindal, D., Gupta, P., Jha, D., Ajay, V. S., Goenka, S., Jacob, P., Mehrotra, K., Perel, P., Nyong, J., Roy, A., Tandon, N., Prabhakaran, D., & Patel, V. (2018). Development of mWellcare: an mHealth intervention for integrated management of hypertension and diabetes in low-resource settings. Glob Health Action, 11(1), 1517930. https://doi.org/10.1080/16549716.2018.1517930 | Outcomes do not address the barriers or facilitators for the adoption and use of telemedicine systems |
| 67 | John, O., & Jha, V. (2019). Remote Patient Management in Peritoneal Dialysis: An Answer to an Unmet Clinical Need. In C. Ronco, C. Crepaldi, & M. H. Rosner (Eds.), REMOTE PATIENT MANAGEMENT IN PERITONEAL DIALYSIS (Vol. 197, pp. 99-112). https://doi.org/10.1159/000496305 | Not focussed on India<br>Outcomes do not address the barriers or facilitators for the adoption and use of telemedicine systems |
| 68 | Kalita, J., Pandey, P. C., Shukla, R., & Misra, U. K. (2021). Feasibility and usefulness of tele-follow-up in the patients with tuberculous meningitis [Article]. Trans R Soc Trop Med Hyg, 115(10), 1153-1159. https://doi.org/10.1093/trstmh/trab069 | Outcomes do not address the barriers or facilitators for the adoption and use of telemedicine systems |
| 69 | Kanchan, S., Saini, L. K., Daga, R., Arora, P., & Gupta, R. (2021). Status of the practice of sleep medicine in India during the COVID-19 pandemic [Article]. J Clin Sleep Med, 17(6), 1229-1235. https://doi.org/10.5664/jcsm.9172 | Outcomes do not address the barriers or facilitators for the adoption and use of telemedicine systems |
| 70 | Kaushik, A., Patel, S., & Dubey, K. (2020). Digital cardiovascular care in COVID-19 pandemic: A potential alternative? [Review]. J Card Surg, 35(12), 3545-3550. https://doi.org/10.1111/jocs.15094 | 3 |

| | | |
|---|---|---|
| 71 | Kavitha, K. V., Deshpande, S. R., Pandit, A. P., & Unnikrishnan, A. G. (2020). Application of tele-podiatry in diabetic foot management: A series of illustrative cases. Diabetes Metab Syndr, 14(6), 1991-1995. https://doi.org/10.1016/j.dsx.2020.10.009 | Outcomes do not address the barriers or facilitators for the adoption and use of telemedicine systems |
| 72 | Kho, J., Gillespie, N., & Martin-Khan, M. (2020). A systematic scoping review of change management practices used for telemedicine service implementations. BMC Health Serv Res, 20(1), 815. https://doi.org/10.1186/s12913-020-05657-w | Not focussed on India |
| 73 | Kodali, P. B., & Das, S. (2021). Digital Health Technologies for Universal Health Coverage: A Promising Change. CURRENT SCIENCE, 120(4), 637-643. https://doi.org/10.18520/cs/v120/i4/637-[illegible] | 1 (ImplementatIon challenges of digital health systems) |
| 74 | Kumar, C. N., Chand, P. K., Manjunatha, N., Math, S. B., Shashidhara, H. N., Basavaraju, V., Thirthalli, J., Manjappa, A. A., Parthasarathy, R., Murthy, P., Ibrahim, F. A., Jagtap, N., Jyrwa, S., Reddy, S., Arora, S., Hawk, M., Kumar, S., Egan, J., & McDonald, M. (2020). Impact Evaluation of VKN-NIMHANS-ECHO Model of Capacity Building for Mental Health and Addiction: Methodology of Two Randomized Controlled Trials. Indian J Psychol Med, 42(6 Suppl), S80-S86. https://doi.org/10.1177/0253717620969066 | Outcomes do not address the barriers or facilitators for the adoption and use of telemedicine systems |
| 75 | Kumar, J., Singh, T., & Singh, A. (2021). Technology connects patients to tertiary care for non-COVID illnesses in pandemic times: A case study from India. INDIAN JOURNAL OF COMMUNITY HEALTH, 33(1), 202-204. https://doi.org/10.47203/IJCH.2020.v33i01.029 | Outcomes do not address the barriers or facilitators for the adoption and use of telemedicine systems |
| 76 | Kumar, M. S., Krishnamurthy, S., Gowda, M. R., & Dhruve, N. (2019). The dawn of eMental health professional. Indian J Psychiatry, 61(Suppl 4), S730-S734. https://doi.org/10.4103/psychiatry.IndianJPsychiatry_161_19 | Not associated with patient health |
| 77 | Kumar, V., Mohanty, P., & Sharma, M. (2021). Promotion of Early Childhood Development Using mHealth: Learnings From an Implementation Experience in Haryana. Indian Pediatr, 58 Suppl 1(SUPPL 1), S37-S41. https://doi.org/10.1007/s13312-021-2354-8 | Outcomes do not address the barriers or facilitators for the adoption and use of telemedicine systems |
| 78 | Lee, M., Kang, D., Yoon, J., Shim, S., Kim, I. R., Oh, D., Shin, S. Y., Hesse, B. W., & Cho, J. (2020). The difference in knowledge and attitudes of using mobile health applications between actual user and non-user among adults aged 50 and older. PLoS One, 15(10), e0241350. https://doi.org/10.1371/journal.pone.0241350 | Not focussed on India |
| 79 | Leochico, C. F. D., & Valera, M. J. S. (2020). Follow-up consultations through telerehabilitation for wheelchair recipients with paraplegia in a developing country: a case report. Spinal Cord Ser Cases, 6(1), 58. https://doi.org/10.1038/s41394-020-0310-9 | Not focussed on India |
| 80 | M, T., & Annamalai, A. (2020). Telepsychiatry and the Role of Artificial Intelligence in Mental Health in Post-COVID-19 India: A Scoping Review on Opportunities [Review]. Indian J Psychol Med, 42(5), 428-434. https://doi.org/10.1177/0253717620952160 | 4 (discusses the scope of AI in telepsychiatry) |

| | | |
|---|---|---|
| 81 | Madanian, S., Parry, D. T., Airehrour, D., & Cherrington, M. (2019). mHealth and big-data integration: promises for healthcare system in India. BMJ Health Care Inform, 26(1). https://doi.org/10.1136/bmjhci-2019-100071 | Outcomes do not address the barriers or facilitators for the adoption and use of telemedicine systems |
| 82 | Mahapatra, P., Sahoo, K. C., Desaraju, S., & Pati, S. (2021). Coping with COVID-19 pandemic: reflections of older couples living alone in urban Odisha, India. Prim Health Care Res Dev, 22, e64. https://doi.org/10.1017/S1463423621000207 | Outcomes do not address the barriers or facilitators for the adoption and use of telemedicine systems |
| 83 | Malathesh, B. C., Ibrahim, F. A., Nirisha, P. L., Kumar, C. N., Chand, P. K., Manjunatha, N., Math, S. B., Thirthalli, J., Manjappa, A. A., Parthasarathy, R., Reddy, S., & Arora, S. (2021). Embracing Technology for Capacity Building in Mental Health: New Path, Newer Challenges. Psychiatr Q, 92(3), 843-850. https://doi.org/10.1007/s11126-020-09859-7 | 1 (staff training) |
| 84 | Maulik, P. K., Kallakuri, S., Devarapalli, S., Vadlamani, V. K., Jha, V., & Patel, A. (2017). Increasing use of mental health services in remote areas using mobile technology: a pre-post evaluation of the SMART Mental Health project in rural India. J Glob Health, 7(1), 010408. https://doi.org/10.7189/jogh.07.010408 | Outcomes do not address the barriers or facilitators for the adoption and use of telemedicine systems |
| 85 | Mehrotra, S., & Tripathi, R. (2018). Recent developments in the use of smartphone interventions for mental health. Curr Opin Psychiatry, 31(5), 379-388. https://doi.org/10.1097/YCO.[illegible] | Not focussed on India |
| 86 | Modi, D., Dholakia, N., Gopalan, R., Venkatraman, S., Dave, K., Shah, S., Desai, G., Qazi, S. A., Sinha, A., Pandey, R. M., Anand, A., Desai, S., & Shah, P. (2019). mHealth intervention "ImTeCHO" to improve delivery of maternal, neonatal, and child care services-A cluster-randomized trial in tribal areas of Gujarat, India. PLoS Med, 16(10), e1002939. https://doi.org/10.1371/journal.pmed.1002939 | 2 (mHealth-based job aid for the ASHAs and PHC staff with an aim to make their work easier and more effective) |
| 87 | Mok, V. C. T., Pendlebury, S., Wong, A., Alladi, S., Au, L., Bath, P. M., Biessels, G. J., Chen, C., Cordonnier, C., Dichgans, M., Dominguez, J., Gorelick, P. B., Kim, S., Kwok, T., Greenberg, S. M., Jia, J., Kalaria, R., Kivipelto, M., Naegandran, K., . . . Scheltens, P. (2020). Tackling challenges in care of Alzheimer's disease and other dementias amid the COVID-19 pandemic, now and in the future [Review]. Alzheimers Dement, 16(11), 1571-1581. https://doi.org/10.1002/alz.12143 | Not focussed on India |
| 88 | Munikrishna, R., Kavitha, T. C., Venkataramanaiah, Somu, G., Kamath, R., D'Souza, B., & Kamath, S. (2019). The Development of a Web Portal for an Assisted Reproduction Center in South India and an Analysis of its Efficacy [Article]. Medico-Legal Update, 19(1), 147-151. https://doi.org/10.5958/0974-1283.2019.00030.6 | Outcomes do not address the barriers or facilitators for the adoption and use of telemedicine systems |
| 89 | Nadhamuni, S., John, O., Kulkarni, M., Nanda, E., Venkatraman, S., Varma, D., Balsari, S., Gudi, N., Samantaray, S., Reddy, H., & Sheel, V. (2021). Driving digital transformation of comprehensive primary health services at scale in India: an enterprise architecture framework. BMJ Glob Health, 6(Suppl 5). https://doi.org/10.1136/bmjgh-2021-005242 | Not associated with patient health |

| | | |
|---|---|---|
| 90 | Narekuli, A., Raja, K., & Pandve, H. T. (2019). Telerehabilitation in India: Points to Ponder. Indian Journal of Physiotherapy and Occupational Therapy - An International Journal, 13(3), 18-21. https://doi.org/10.5958/0973-5674.2019.00084.4 | Outcomes do not address the barriers or facilitators for the adoption and use of telemedicine systems |
| 91 | Naslund, J. A., Aschbrenner, K. A., Araya, R., Marsch, L. A., Unutzer, J., Patel, V., & Bartels, S. J. (2017). Digital technology for treating and preventing mental disorders in low-income and middle-income countries: a narrative review of the literature. Lancet Psychiatry, 4(6), 486-500. https://doi.org/10.1016/S2215-0366(17)30096-2 | Not focussed on India |
| 92 | Opoku, D., Stephani, V., & Quentin, W. (2017). A realist review of mobile phone-based health interventions for non-communicable disease management in sub-Saharan Africa. BMC Med, 15(1), 24. https://doi.org/10.1186/s12916-017-0782-z | Not focussed on India |
| 93 | Pai, R. R., & Alathur, S. (2020). Mobile health intervention and COVID-19 pandemic outbreak: insights from Indian context. International Journal of Health Governance, 26(1), 42-50. https://doi.org/10.1108/ijhg-04-2020-0043 | 4 (mHealth intervention for COVID-19) |
| 94 | Pasquali, P., Sonthalia, S., Moreno-Ramirez, D., Sharma, P., Agrawal, M., Gupta, S., Kumar, D., & Arora, D. (2020). Teledermatology and its Current Perspective. Indian Dermatol Online J, 11(1), 12-20. https://doi.org/10.4103/idoj.IDOJ_241_19 | 3 |
| 95 | Patel, A., Praveen, D., Maharani, A., Oceandy, D., Pilard, Q., Kohli, M. P. S., Sujarwoto, S., & Tampubolon, G. (2019). Association of Multifaceted Mobile Technology-Enabled Primary Care Intervention With Cardiovascular Disease Risk Management in Rural Indonesia. JAMA Cardiol, 4(10), 978-986. https://doi.org/10.1001/jamacardio.2019.2974 | Not focussed on India |
| 96 | Patel, S. A., Vashist, K., Jarhyan, P., Sharma, H., Gupta, P., Jindal, D., Venkateshmurthy, N. S., Pfadenhauer, L., Mohan, S., & Tandon, N. (2021). A model for national assessment of barriers for implementing digital technology interventions to improve hypertension management in the public health care system in India. BMC Health Serv Res, 21(1), 1101. https://doi.org/10.1186/s12913-021-06999-9 | 1 (evaluated existing healthcare infrastructure to support digital health interventions and examined epidemiologic, socioeconomic, and geographical contexts) |
| 97 | Polanco, E., Aquey, M., Collado, J., Campos, E., Guzman, J., Cuevas-Budhart, M. A., Divino-Filho, J. C., & Ramos-Sanchez, A. (2021). A COVID-19 pandemic-specific, structured care process for peritoneal dialysis patients facilitated by telemedicine: Therapy continuity, prevention, and complications management. Ther Apher Dial, 25(6), 970-978. https://doi.org/10.1111/1744-9987.13635 | Not focussed on India |
| 98 | Pradeepa, R., Rajalakshmi, R., & Mohan, V. (2019). Use of Telemedicine Technologies in Diabetes Prevention and Control in Resource-Constrained Settings: Lessons Learned from Emerging Economies [Article]. Diabetes Technol Ther, 21(S2), S29-S216. https://doi.org/10.1089/dia.2019.0038 | 3, 4 |

| | | |
|---|---|---|
| 99 | Ramalho, R., Adiukwu, F., Gashi Bytyci, D., El Hayek, S., Gonzalez-Diaz, J. M., Larnaout, A., Grandinetti, P., Nofal, M., Pereira-Sanchez, V., Pinto da Costa, M., Ransing, R., Teixeira, A. L. S., Shalbafan, M., Soler-Vidal, J., Syarif, Z., & Orsolini, L. (2020). Telepsychiatry During the COVID-19 Pandemic: Development of a Protocol for Telemental Health Care [Article]. Front Psychiatry, 11, 552450, Article 552450. https://doi.org/10.3389/fpsyt.2020.552450 | Not focussed on India<br>Outcomes do not address the barriers or facilitators for the adoption and use of telemedicine systems |
| 100 | Rathi, V. M., Das, A. V., & Khanna, R. C. (2020). Impact of COVID-19-related lockdown-I on a network of rural eye centres in Southern India [Article]. Indian J Ophthalmol, 68(11), 2396-2398. https://doi.org/10.4103/ijo.IJO_2303_20 | Outcomes do not address the barriers or facilitators for the adoption and use of telemedicine systems |
| 101 | Ravi, R., Gunjawate, D. R., Yerraguntla, K., & Driscoll, C. (2018). Knowledge and Perceptions of Teleaudiology Among Audiologists: A Systematic Review. J Audiol Otol, 22(3), 120-127. https://doi.org/10.7874/jao.2017.00353 | Not focussed on India |
| 102 | Ravindran, S., P, L. N., Channaveerachari, N. K., Seshadri, S. P., Kasi, S., Manikappa, S. K., Cherian, A. V., Palanimuthu, T. S., Sudhir, P., Govindan, R., P, B. R., Christopher, A. D., & George, S. (2020). Crossing barriers: Role of a tele-outreach program addressing psychosocial needs in the midst of COVID-19 pandemic. Asian J Psychiatr, 53, 102351. https://doi.org/10.1016/j.ajp.2020.102351 | Outcomes do not address the barriers or facilitators for the adoption and use of telemedicine systems |
| 103 | Rout, S. K., Gabhale, Y. R., Dutta, A., Balakrishnan, S., Lala, M. M., Setia, M. S., Bhuyan, K., & Manglani, M. V. (2019). Can telemedicine initiative be an effective intervention strategy for improving treatment compliance for pediatric HIV patients: Evidences on costs and improvement in treatment compliance from Maharashtra, India. PLoS One, 14(10), e0223303. https://doi.org/10.1371/journal.pone.0223303 | Outcomes do not address the barriers or facilitators for the adoption and use of telemedicine systems |
| 104 | Sarveswaran, G., Rangamani, S., Ghosh, A., Bhansali, A., Dharmalingam, M., Unnikrishnan, A. G., Kishore Vikram, N., Mathur, P., & Misra, A. (2021). Management of diabetes mellitus through teleconsultation during COVID-19 and similar scenarios - Guidelines from Indian Council of Medical Research (ICMR) expert group. Diabetes Metab Syndr, 15(5), 102242. https://doi.org/10.1016/j.dsx.2021.102242 | Not concerned with patient monitoring, screening, diagnostic tests, or treatment |
| 105 | Sawadkar, M. M., & Nayak, V. R. (2021). Telehealth: The role of respiratory therapists during the COVID-19 emergency [Letter]. Can J Respir Ther, 57, 119-120. https://doi.org/10.29390/cjrt-2021-[illegible] | Not concerned with patient monitoring, screening, diagnostic tests, or treatment |
| 106 | Seethalakshmi, S., & Nandan, R. (2020). Health is the Motive and Digital is the Instrument. J Indian Inst Sci, 100(4), 1-6. https://doi.org/10.1007/s41745-020-00190-5 | Outcomes do not address the barriers or facilitators for the adoption and use of telemedicine systems |
| 107 | Shah, P., Madhiwala, N., Shah, S., Desai, G., Dave, K., Dholakia, N., Patel, S., Desai, S., & Modi, D. (2019). High uptake of an innovative mobile phone application among community health workers in rural India: An implementation study. Natl Med J India, 32(5), 262-269. https://doi.org/10.4103/0970-258X.295956 | Not concerned with patient monitoring, screening, diagnostic tests, or treatment |

| | | |
|---|---|---|
| 108 | Shah, P., Yauney, G., Gupta, O., Patalano II, V., Mohit, M., Merchant, R., & Subramanian, S. V. (2018). Technology-enabled examinations of cardiac rhythm, optic nerve, oral health, tympanic membrane, gait and coordination evaluated jointly with routine health screenings: an observational study at the 2015 Kumbh Mela in India. BMJ Open, 8(4), e018774. https://doi.org/10.1136/bmjopen-2017-018774 | Outcomes do not address the barriers or facilitators for the adoption and use of telemedicine systems |
| 109 | Sharma, H., Suprabha, B. S., & Rao, A. (2021). Teledentistry and its applications in paediatric dentistry: A literature review [Review]. Pediatr Dent J, 31(3), 203-215. https://doi.org/10.1016/j.pdj.2021.08.003 | 3, 4 (barriers and facilitators are dicussed in the event of a pandemic & for this intervention) |
| 110 | Sikka, V., Somma, S. D., Galwankar, S. C., Sinha, S., Garg, N., Talwalkar, N., Garg, S., Mahajan, P., Chauhan, V., Moreno-Walton, L., Dubhashi, S., Dutta, V., Saddikuti, V., PW, B. N., Grover, J., Paranjape, K., Singh, S., Sharma, P., Bhoi, S., . . . Sardesai, I. (2021). The World Health Organization Collaborating Center for Emergency and Trauma (WHO-CCET) in South East Asia, The World Academic Council of Emergency Medicine (WACEM), and The American College of Academic International Medicine (ACAIM) 2021 Framework for using Telemedicine Technology at Healthcare Institutions [Article]. J Emerg Trauma Shock, 14(3), 173-179. https://doi.org/10.4103/jets.jets_105_21 | Not focussed on India |
| 111 | Singh Pardal, M., Rajiva, K., & Orkeh, G. (2020). Telemedicine in the era of COVID-19: The East and the West. JOURNAL OF MARINE MEDICAL SOCIETY, 0(0), 32-35. https://doi.org/10.4103/jmms.jmms_86_20 | 3 |
| 112 | Singhal, S., Das, S., Dubey, S., Sahu, M. K., Kumar, M., & Dubey, R. K. (2020). Technology- Can it Emancipate the Void in India's Mental Healthcare Delivery? Journal of Evolution of Medical and Dental Sciences, 9(06), 335-338. https://doi.org/10.14260/jemds/2020/76 | 4 (discusses the scope for developing a new mHealth intervention in the future for mental health awareness of caregivers; barriers and facilitators are not discussed) |
| 113 | Sinha Deb, K., Tuli, A., Sood, M., Chadda, R., Verma, R., Kumar, S., Ganesh, R., & Singh, P. (2018). Is India ready for mental health apps (MHApps)? A quantitative-qualitative exploration of caregivers' perspective on smartphone-based solutions for managing severe mental illnesses in low resource settings. PLoS One, 13(9), e0203353. https://doi.org/10.1371/journal.pone.0203353 | 4 (discusses the scope scope for deploying smartphone based solutions to improve illness management and reduce caregiver burden) |
| 114 | Sivakumar, P. T., Mukku, S. S. R., Kar, N., Manjunatha, N., Phutane, V. H., Sinha, P., Kumar, C. N., & Math, S. B. (2020). Geriatric Telepsychiatry: Promoting Access to Geriatric Mental Health Care Beyond the Physical Barriers. Indian J Psychol Med, 42(5 Suppl), 41S-46S. https://doi.org/10.1177/0253717620958380 | 4 |
| 115 | Sreejith, G., & Menon, V. (2019). Mobile Phones as a Medium of Mental Health Care Service Delivery: Perspectives and Barriers among Patients with Severe Mental Illness. Indian J Psychol Med, 41(5), 428-433. https://doi.org/10.4103/IJPSYM.IJPSYM_333_18 | 4 (discusses the user perspectives and service delivery preferences before the implementation of mobile phone based interventions) |

| | | |
|---|---|---|
| 116 | Srinivasan, R., Wallis, K. E., & Soares, N. (2022). Global Trends in Telehealth Among Clinicians in Developmental-Behavioral Pediatric Practice: A COVID-19 Snapshot. J Dev Behav Pediatr, 43(1), 32-37. https://doi.org/10.1097/DBP.0000000000000963 | 3 |
| 117 | Srivastava, A., Swaminathan, A., Chockalingam, M., Srinivasan, M. K., Surya, N., Ray, P., Hegde, P. S., Akkunje, P. S., Kamble, S., Chitnis, S., Kamalakannan, S., Ganvir, S., Shah, U., & Indian Federation of Neurorehabilitation Research Task, F. (2021). Tele-Neurorehabilitation During the COVID-19 Pandemic: Implications for Practice in Low- and Middle-Income Countries [Article]. Front Neurol, 12, 667925, Article 667925. https://doi.org/10.3389/fneur.2021.667925 | Not focussed on India |
| 118 | Subathra, G. N., Rajendrababu, S. R., Senthilkumar, V. A., Mani, I., & Udayakumar, B. (2021). Impact of COVID-19 on follow-up and medication adherence in patients with glaucoma in a tertiary eye care centre in south India [Article]. Indian J Ophthalmol, 69(5), 1264-1270. https://doi.org/10.4103/ijo.IJO_164_21 | Outcomes do not address the barriers or facilitators for the adoption and use of telemedicine systems |
| 119 | Suresh, L. R., & Hegde, A. M. (2021). Feasibility of teledentistry in population groups: Introducing a matrix model for its assessment. WORLD MEDICAL & HEALTH POLICY, 13(4), 758-765. https://doi.org/10.1002/wmh3.473 | 1,4 (describes a system to facilitate transition to teledentistry by assessing the feasibility based on target population characteristics) |
| 120 | Suryavanshi, N., Kadam, A., Kanade, S., Gupte, N., Gupta, A., Bollinger, R., Mave, V., & Shankar, A. (2020). Acceptability and feasibility of a behavioral and mobile health intervention (COMBIND) shown to increase uptake of prevention of mother to child transmission (PMTCT) care in India. BMC Public Health, 20(1), 752. https://doi.org/10.1186/s12889-020-08706-5 | Outcomes do not address the barriers or facilitators for the adoption and use of telemedicine systems |
| 121 | Talati, K., Amin, A. A., & Nimbalkar, S. M. (2018). Implementation of a Pilot Test mHealth Application to Improve Home Based Newborn Care (IMNCI) in Remote Tribal Gujarat. JOURNAL OF CLINICAL AND DIAGNOSTIC RESEARCH, 12(6), SE01-SE03. https://doi.org/10.7860/jcdr/2018/35229.11687 | Not concerned with patient monitoring, screening, diagnostic tests, or treatment<br>Outcomes do not address the barriers or facilitators for the adoption and use of telemedicine systems |
| 122 | Tewari, A., Kallakuri, S., Devarapalli, S., Jha, V., Patel, A., & Maulik, P. K. (2017). Process evaluation of the systematic medical appraisal, referral and treatment (SMART) mental health project in rural India. BMC Psychiatry, 17(1), 385. https://doi.org/10.1186/s12888-017-1525-6 | Outcomes do not address the barriers or facilitators for the adoption and use of telemedicine systems |
| 123 | Tewari, A., Kallakuri, S., Devarapalli, S., Peiris, D., Patel, A., & Maulik, P. K. (2021). SMART Mental Health Project: process evaluation to understand the barriers and facilitators for implementation of multifaceted intervention in rural India. Int J Ment Health Syst, 15(1), 15. https://doi.org/10.1186/s13033-021-00438-2 | Outcomes do not address the barriers or facilitators for the adoption and use of telemedicine systems |
| 124 | Ummer, O., Scott, K., Mohan, D., Chakraborty, A., & LeFevre, A. E. (2021). Connecting the dots: Kerala's use of digital technology during the COVID-19 response. BMJ Glob Health, 6(Suppl 5). https://doi.org/10.1136/bmjgh-2021-005355 | Outcomes do not address the barriers or facilitators for the adoption and use of telemedicine systems |

| | | |
|---|---|---|
| 125 | Upadhyayula, P. S., Yue, J. K., Yang, J., Birk, H. S., & Ciacci, J. D. (2018). The Current State of Rural Neurosurgical Practice: An International Perspective. J Neurosci Rural Pract, 9(1), 123-131. https://doi.org/10.4103/jnrp.jnrp_273_17 | Not focussed on India |
| 126 | Usmanova, G., Gresh, A., Cohen, M. A., Kim, Y. M., Srivastava, A., Joshi, C. S., Bhatt, D. C., Haws, R., Wadhwa, R., Sridhar, P., Bahl, N., Gaikwad, P., & Anderson, J. (2020). Acceptability and Barriers to Use of the ASMAN Provider-Facing Electronic Platform for Peripartum Care in Public Facilities in Madhya Pradesh and Rajasthan, India: A Qualitative Study Using the Technology Acceptance Model-3. Int J Environ Res Public Health, 17(22). https://doi.org/10.3390/ijerph17228333 | 2 |
| 127 | van der Watt, A. S. J., Odendaal, W., Louw, K., & Seedat, S. (2020). Distant mood monitoring for depressive and bipolar disorders: a systematic review. BMC Psychiatry, 20(1), 383. https://doi.org/10.1186/s12888-020-02782-y | Not focussed on India |
| 128 | Vinayagamoorthy, K., Acharya, S., Kumar, M., Pentapati, K. C., & Acharya, S. (2019). Efficacy of a remote screening model for oral potentially malignant disorders using a free messaging application: A diagnostic test for accuracy study [Article]. Aust J Rural Health, 27(2), 170-176. https://doi.org/10.1111/ajr.12496 | Outcomes do not address the barriers or facilitators for the adoption and use of telemedicine systems |
| 129 | Yadav, S., Sethi, R., Pradhan, A., Vishwakarma, P., Bhandari, M., Gattani, R., Chandra, S., Chaudhary, G., Sharma, A., Dwivedi, S. K., Narain, V. S., Rao, B., & Roy, A. (2021). 'Routine' versus 'Smart Phone Application Based - Intense' follow up of patients with acute coronary syndrome undergoing percutaneous coronary intervention: Impact on clinical outcomes and patient satisfaction [Article]. Int J Cardiol Heart Vasc, 35, 100832, Article 100832. https://doi.org/10.1016/j.ijcha.2021.100832 | Outcomes do not address the barriers or facilitators for the adoption and use of telemedicine systems |
| 130 | Yeoh, E., Tan, S. G., Lee, Y. S., Tan, H. H., Low, Y. Y., Lim, S. C., Sum, C. F., Tavintharan, S., & Wee, H. L. (2021). Impact of COVID-19 and partial lockdown on access to care, self-management and psychological well-being among people with diabetes: A cross-sectional study. Int J Clin Pract, 75(8), e14319. https://doi.org/10.1111/ijcp.14319 | Not focussed on India |
| 131 | Yim, D., Chandra, S., Sondh, R., Thottarath, S., & Sivaprasad, S. (2021). Barriers in establishing systematic diabetic retinopathy screening through telemedicine in low- and middle-income countries. Indian J Ophthalmol, 69(11), 2987-2992. https://doi.org/10.4103/ijo.IJO_1411_21 | Not focussed on India |
